# Investigation of the genetic architecture of cam morphology, and its relationship with hip osteoarthritis, using alpha angle as a proxy measure

**DOI:** 10.1101/2022.07.22.22277884

**Authors:** Benjamin G. Faber, Monika Frysz, April E. Hartley, Raja Ebsim, Cindy G. Boer, Fiona R. Saunders, Jennifer S. Gregory, Richard M Aspden, Nicholas C. Harvey, Lorraine Southam, William Giles, Christine Le Maitre, J. Mark Wilkinson, Joyce B.J. van Meurs, Eleftheria Zeggini, Timothy Cootes, Claudia Lindner, John P. Kemp, George Davey Smith, Jonathan H. Tobias

## Abstract

**Objectives:** To examine the genetic architecture of cam morphology, using alpha angle (AA) as a proxy measure, we conducted an AA genome wide association study (GWAS), followed by Mendelian randomisation (MR) to evaluate its causal relationship with hip osteoarthritis (HOA).

**Methods:** Observational analyses examined associations between AA derived from hip DXA scans in UK Biobank (UKB), and radiographic HOA (rHOA) and subsequent total hip replacement (THR). Afterwards, an AA GWAS meta-analysis was performed (n=44,214), using AA previously derived in the Rotterdam Study (RS). Linkage disequilibrium score regression assessed the genetic correlation between AA and HOA. Genetic associations with P<5×10^−8^ instrumented AA for two-sample MR.

**Results:** DXA-derived AA showed expected associations between AA and rHOA (OR 1.63 [95% CI 1.58-1.67]) and THR (HR 1.45 [1.33-1.59]) in UKB. The heritability of AA was 10% and AA had a moderate genetic correlation with HOA (r_g_=0.26 [0.10-0.43]). Eight independent genetic signals were associated with AA. Two-sample MR provided weak evidence of causal effects of AA on HOA risk (inverse variance weighted (IVW): OR=1.84 [1.14-2.96], P 0.01). In contrast, genetic predisposition for HOA had stronger evidence of a causal effect on increased AA (IVW: β=0.09 [0.04-0.13], P 4.58 × 10^−05^).

**Conclusions:** Expected observational associations between AA and related clinical outcomes provided face-validity for the DXA-derived AA measures. Evidence of bidirectional associations between AA and HOA, particularly in the reverse direction, suggest that hip shape remodelling secondary to a genetic predisposition to HOA contribute to the well-established relationship between HOA and cam morphology in older adults.

## Introduction

Cam morphology describes a non-spherical femoral head which has been associated with hip osteoarthritis (HOA) (1, 2). Longitudinal studies have shown cam morphology precedes HOA and from this causation has been inferred (2, 3), prompting research into the benefits of surgical correction (4, 5). That said, observational studies showing temporal associations still suffer from confounding making causal inferences difficult (1, 6, 7).

Alpha angle (AA), a measure of femoral head sphericity, is widely used to define cam morphology with a higher angle considered to be more severe (4, 8). Cam morphology has been suggested to develop in adolescence due to antero-lateral femoral head offset or increased impact as the growth plate fuses leading to greater bone deposition (9, 10). Additionally, a similar morphology may develop in later life as a consequence of modelling changes occurring as part of the osteoarthritic process. Croft scoring for HOA specifically recognises abnormal hip shape as the last stage of osteoarthritis (OA) (11).

Femoro-acetabular impingement (FAI) has been proposed to explain the causal pathway between cam morphology and HOA (12). FAI syndrome encompasses individuals with hip pain, coexistent with cam morphology and specific examination findings (13). FAI syndrome is seen predominantly in younger adults before the onset of HOA. Surgical interventions to remove cam lesions in this population have been evaluated with limited success (4, 5). As well as potentially improving hip pain, these surgical procedures have been suggested to prevent the development or progression of HOA (5).

One method to derive causal inferences from observational data is Mendelian randomisation (MR) which uses genetic loci as instrumental variables and largely removes the effects of confounding and reverse causation (14). The UK Biobank study (UKB), a cohort study of adults aged 40-69 years at inception, provides the sample size required to study the relationship between hip shape and HOA using MR (15, 16). In this study, we aimed to provide face validity for our novel automated method for deriving AA from hip DXA scans in UKB by confirming expected relationships with HOA, perform an AA GWAS meta-analysis to establish the genetic architecture of cam morphology, and finally use MR analysis based on genetic instruments identified from our AA GWAS to establish whether there is a causal relationship between increased AA and HOA.

## Patients & Methods

### Alpha Angle

This study included UKB participants with a left hip dual-energy X-ray absorptiometry (DXA) scan (iDXA GE-Lunar, Madison, WI). Outline points were automatically placed around each hip and features of radiographic HOA (rHOA) were measured semi automatically as previously described (15, 17, 18). AA was estimated using the outline points that excluded osteophytes (Figure 1) and a previously published Python code (19, 20). This study also included individuals from the Rotterdam Study (RS) who had AA measured from anterior-posterior pelvic radiographs using similar methods (see Supplementary Methods) (21). Ethics approval was given by the appropriate body for each study (see Ethics approval statement).

**Figure 1.**
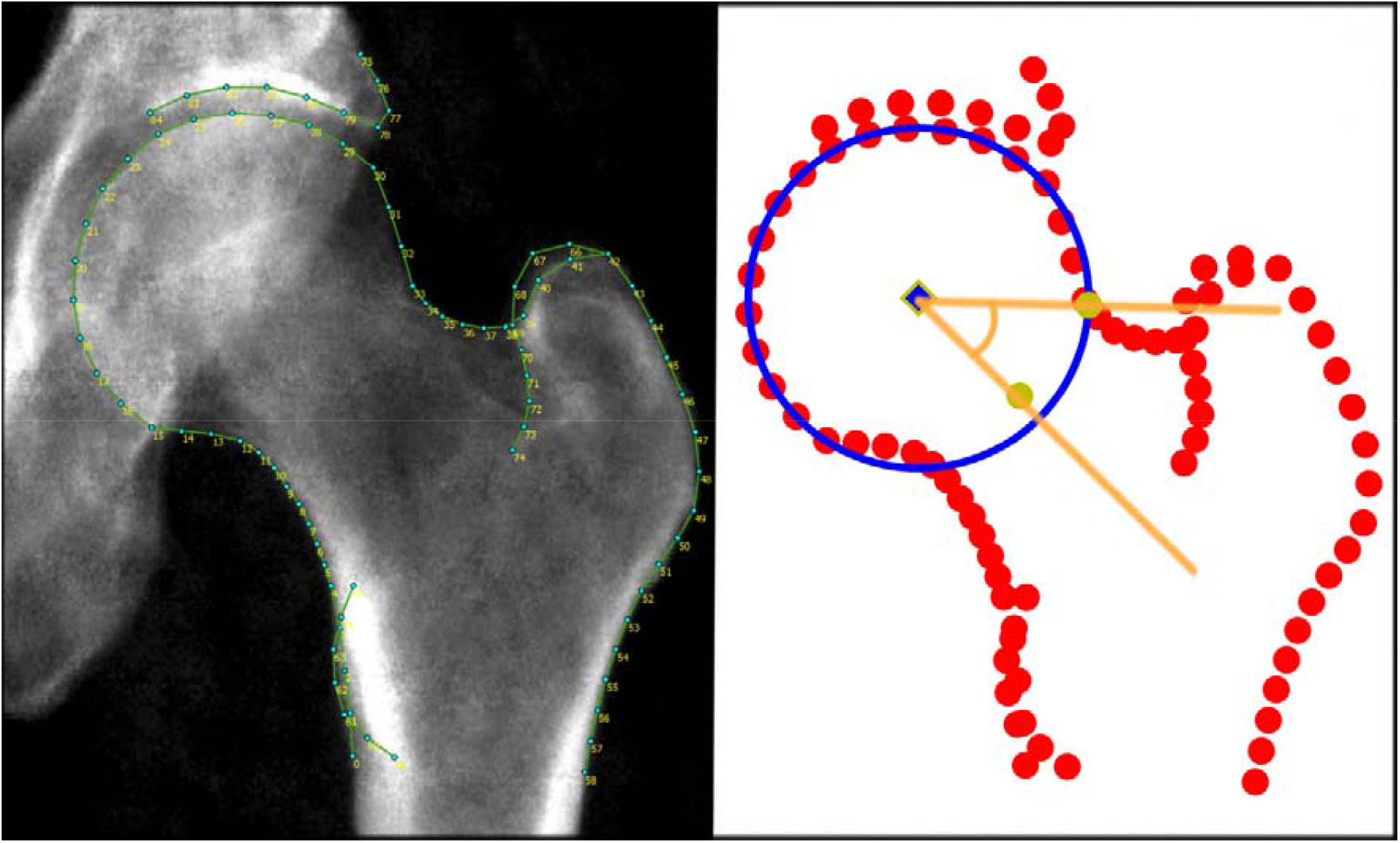
Calculating alpha angle automatically in UK Biobank. Left – UK Biobank DXA image with outline points marked and lines connecting the points. Right - the same points are visualised in Python, where a circle of best fit is plotted, and the AA is calculated from the femoral neck mid-point (yellow) and the point at which the femoral neck intersects the circle (yellow). In this individual the AA is 41.7°.

### Outcome measures of osteoarthritis and observational associations in UK Biobank

UKB participants were asked whether they had hip pain for >3months via questionnaire on the same day as their DXA scan. Hospital diagnosed HOA was based on hospital episode statistics (HES) data, termed HES OA, as was total hip replacement (THR) (17). Logistic regression was used to examine associations between AA with clinical outcomes, apart from with THR which was examined using Cox proportional hazard modelling. Further sensitivity analyses were performed defining cam morphology as an AA ≥60° (see Supplementary Methods).

### Alpha angle genome-wide association study

AA was standardised to create a Z-score (standard deviation (SD)=1, mean =0). Subsequently, a GWAS meta-analysis of AA was conducted between a GWAS in each study (UKB n=38,173, RS1 (n=2,970), RSII (n=1,817) & RSIII (n=1,254)). EasyQC was used to clean and harmonise the data (22), SNPs with a minor allele frequency (MAF) <0.01 and imputation score <0.4 were removed. Meta-analysis between the studies was performed using fixed-effects inverse variance weighting with METAL (23). A threshold of P-value <5×10^−8^ was used to define genome-wide significance. The independent SNPs of interest for AA were identified using linkage disequilibrium (LD) clumping (see MR methods section). A sensitivity analysis using GCTA-COJO to verify independent SNPs was also done (24). Genetic correlations and heritability were estimated using LD score regression (LDSR) (25). See Supplementary Methods for further details.

### Downstream analyses

Expression quantitative trait loci (eQTL) database GTEx v8 was searched for each SNP to identify cis-acting effects (26). Bayesian colocalisation was used to identify cis-acting genes within GTEx, in tissues with the greatest evidence of expression and cultured cell fibroblast given their similarity to joint tissue, using LocusFocus (27). In addition, colocalisation methods were utilised to examine eQTL data taken from human cartilage, a tissue that is not readily available in eQTL databases, using the coloc.fast package in R (28, 29). One megabase was examined either side of the sentinel SNP. Details of the cartilage samples can be found here (29) but briefly eQTL data were assessed on highly (diseased) and less degraded (healthy) cartilage retrieved following knee and hip joint replacements. A SNP was considered to colocalise with an eQTL if the posterior probability (PP) was >80% and suggestive if PP >60% (28). Regulatory elements of non-coding human genome were identified using RegulomeDB (30). Finally, immunohistochemistry staining in human knee osteochondral tissue was examined for any gene which colocalised in human cartilage (Supplementary Methods).

### Mendelian randomisation

Our AA GWAS meta-analysis provided genetic instruments for AA in two-sample MR. SNPs at p<5×10^−8^ were selected to satisfy the relevance assumption (31), having removed those which were palindromic with a minor allele frequency (MAF) >0.42, or those with a MAF<0.01. LD clumping was applied, using the TwoSampleMR package in R (32), to identify independent genetic signals by removing SNPs that are in LD (r^2^>0.001). A similar approach was used to provide a genetic instrument for HOA based on a GWAS of HES OA in UKB, for use in bi-directional MR analyses. This HOA GWAS excluded all individuals who had had a hip DXA to prevent any sample overlap with the AA GWAS. The MR analyses used the UKB AA GWAS rather than the meta-analysis to reduce heterogeneity. Steiger filtering was applied to strengthen evidence that genetic instruments were upstream of the outcome (32). Bi-directional two sample MR was conducted using the inverse variance weighted method. Sensitivity analyses were conducted that are more robust to violations of the independence and exclusion restriction assumptions, they included MR Egger, weighted median, simple mode and weighted mode, along with single SNP and leave one out analyses (16). Further, we used causal analysis using summary effect estimates (MR-CAUSE) to examine for evidence of causality whilst considering the effects of both correlated and uncorrelated horizontal pleiotropy (33). The MR-STROBE guidelines provided a framework for this analysis (31).

## Results

### Observational associations

In order to provide validity for our DXA measurement of AA, the cross-sectional association of AA with clinical HOA outcomes was examined in our UKB population (n=40,337, Supplementary Table 1). The mean AA was 47.8° (31.8-115.0) with a positively skewed distribution (Supplementary Figure 1) similar to a previous study (34). In both unadjusted and adjusted analyses, higher AA was associated with hip pain (adjusted results: OR 1.15 [95% CI 1.11-1.19]), rHOA grade ≥2 (1.63 [1.58-1.67]), HES OA (1.44 [1.35-1.54]) and subsequent THR (HR 1.45 [1.33-1.59]) (Table 1). Similar associations were seen when investigating cam morphology as a binary variable (Supplementary Table 2).

**Table 1.**
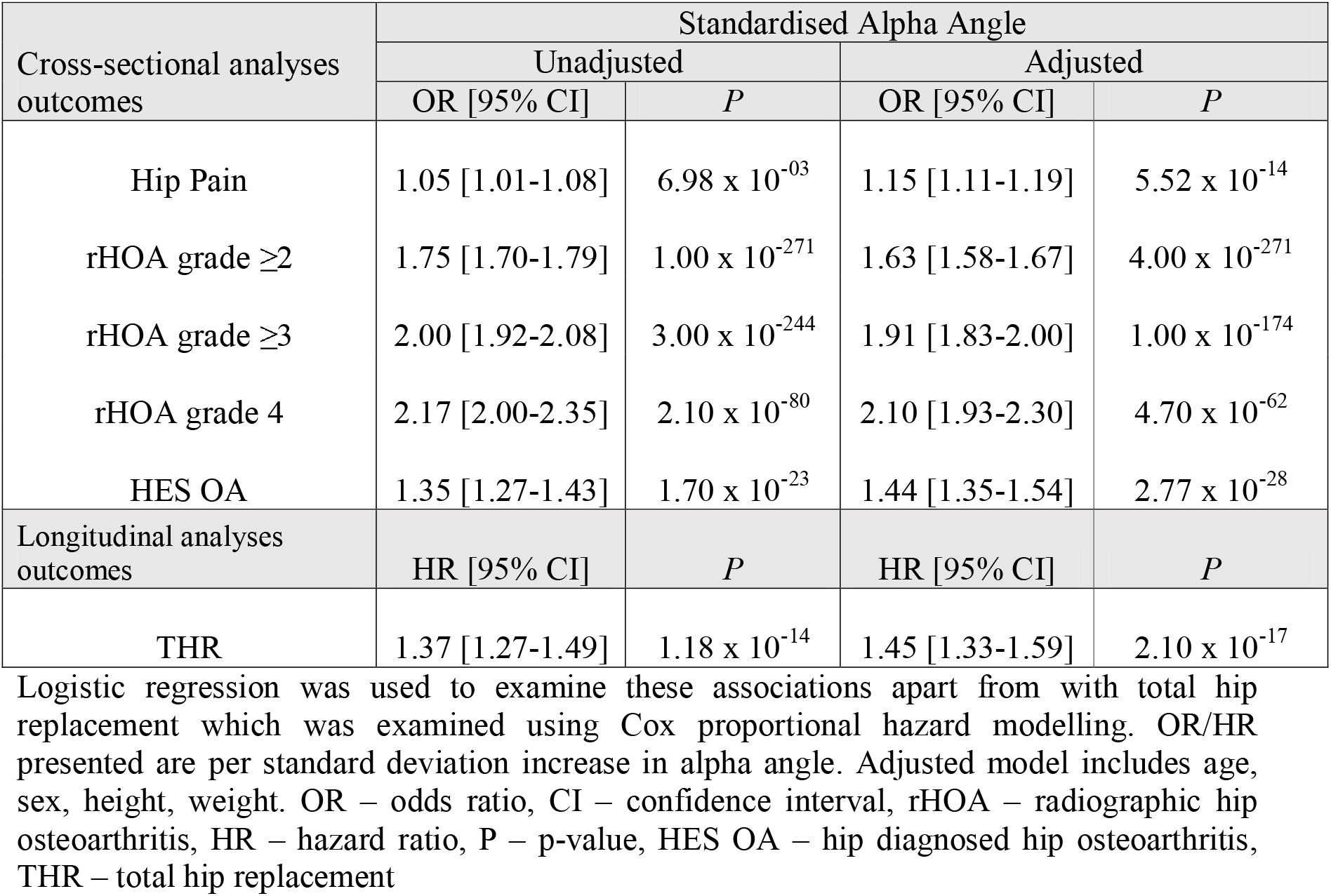
Cross-sectional and longitudinal associations between standardised alpha angle and osteoarthritis outcomes in UK Biobank.

### Alpha Angle GWAS

The GWAS meta-analysis comprised 44,214 participants (Supplementary Figure 2). The Manhattan plot showed 8 genome-wide significant signals (Supplementary Figure 3). The QQ plot showed some genetic inflation (λ 1.08) which was expected (Supplementary Figure 4). SNP trait heritability was modest (h^2^ 0.10). After LD clumping, 8 independent SNPs remained from the meta-analysis, see Supplementary Figure 5 for locus zoom plots of these signals (Table 2). Rs561578905 was the only genome-wide significant hit after meta-analysis that was not present in RS. To mitigate the effects of this, the SNP in highest LD (rs7302982, r^2^ 0.77) was used instead for meta-analysis in the RS. Three SNPs showed weak evidence of heterogeneity (rs7571789: I^2^ 53, heterogeneity P 0.09; rs10478422 I^2^ 33, P 0.21; rs561578905 I^2^ 25, P 0.26) (Supplementary Figure 6). The lead 8 SNPs showed the same direction of effect in a GWAS of cam morphology (AA ≥60°, n=38,173 in UKB) albeit with p-values above our genome-wide significance threshold (Supplementary Table 3). Seven independent SNPs were identified by GCTA-COJO (Supplementary Table 4), six were the same SNPs as those identified by LD clumping, rs561578905 (*SOX5*) was removed and rs10478422 was replaced by rs455991 (*TNFAIP8*, r^2^ 0.97). The closest gene to each independent SNP associated with AA was initially used to label the loci, namely *TGFA, TNFAIP8, TFB1M-TIAM2, LMX1B, GRK5, SOX5, CYP19A1* and *UQCC1*) (Table 2). *TGFA, LMX1B, SOX5, CYP19A1* and *UQCC1-GDF5* loci have previously been associated with OA (35). The *LMX1B* locus shared the same sentinel SNP for AA and OA whereas *SOX5, TGFA, UQCC1-GDF5* AA SNPs were in moderate-high LD (r^2^ 0.30, 0.65 & 0.79 respectively) and *CYP19AI* showed only very weak LD (r^2^ 0.06) with their OA equivalents.

**Table 2.**
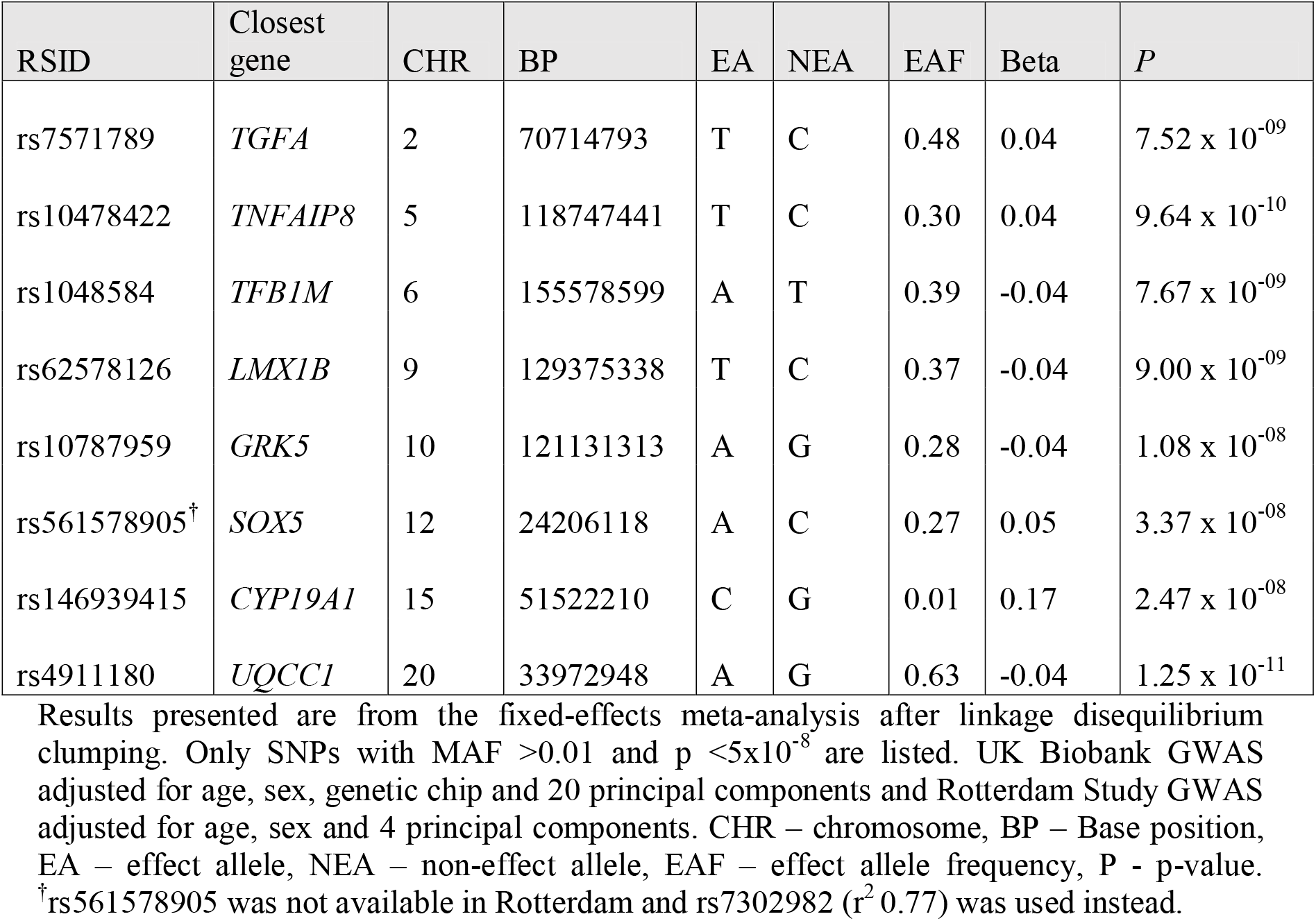
The top independent single nucleotide polymorphisms associated with alpha angle.

#### Downstream analysis of alpha angle hits

The GTEx consortium eQTL database suggested rs10478422, rs62578126 and rs1048584 were cis-eQTLs for *TNFAIP8* (cultured fibroblasts, PP 0.97), *LMX1B* (adipose tissue, PP 0.96), and *CLDN20*/*RP11-477D19*/*TFB1M* (cultured fibroblasts, PP 0.87/0.90/0.66), as they showed evidence of colocalisation (Supplementary Table 5). In further eQTL studies based on human cartilage (n=115), the AA genetic association signal at the *TNFAIP8* locus colocalized with the cis-eQTL signal in highly degraded human cartilage (PP 0.97, Supplementary Figure 7). No other SNPs showed evidence of colocalisation with eQTL data from less or highly degraded cartilage (Supplementary Table 6). Rs7571789 (*TGFA*), rs6595186 (*TNFAIP8)*, rs62578126 (*LMX1B)*, rs561578905 (*SOX5)* and rs246939415 (*CYP19A1)* were all predicted to affect enhancer or promotor activity, based on RegulomeDB probability scores >0.5 (Supplementary Table 7).

Given the finding that the *TNFAIP8* locus colocalized with the cis-eQTL signal in highly degraded human cartilage, we used immunohistochemistry to further explore *TNFAIP8* expression in human knee cartilage and bone (n=4). TNFAIP8 immunopositivity was localised to chondrocytes and osteocytes within the osteochondral tissue samples (Figure 2-A). An increase in percentage immunopositivity in both chondrocytes (P=5.1×10^−3^) and osteocytes (P=2.5 ×10^−3^) was seen in highly degraded compared to less degraded tissues (Figure 2-B).

**Figure 2.**
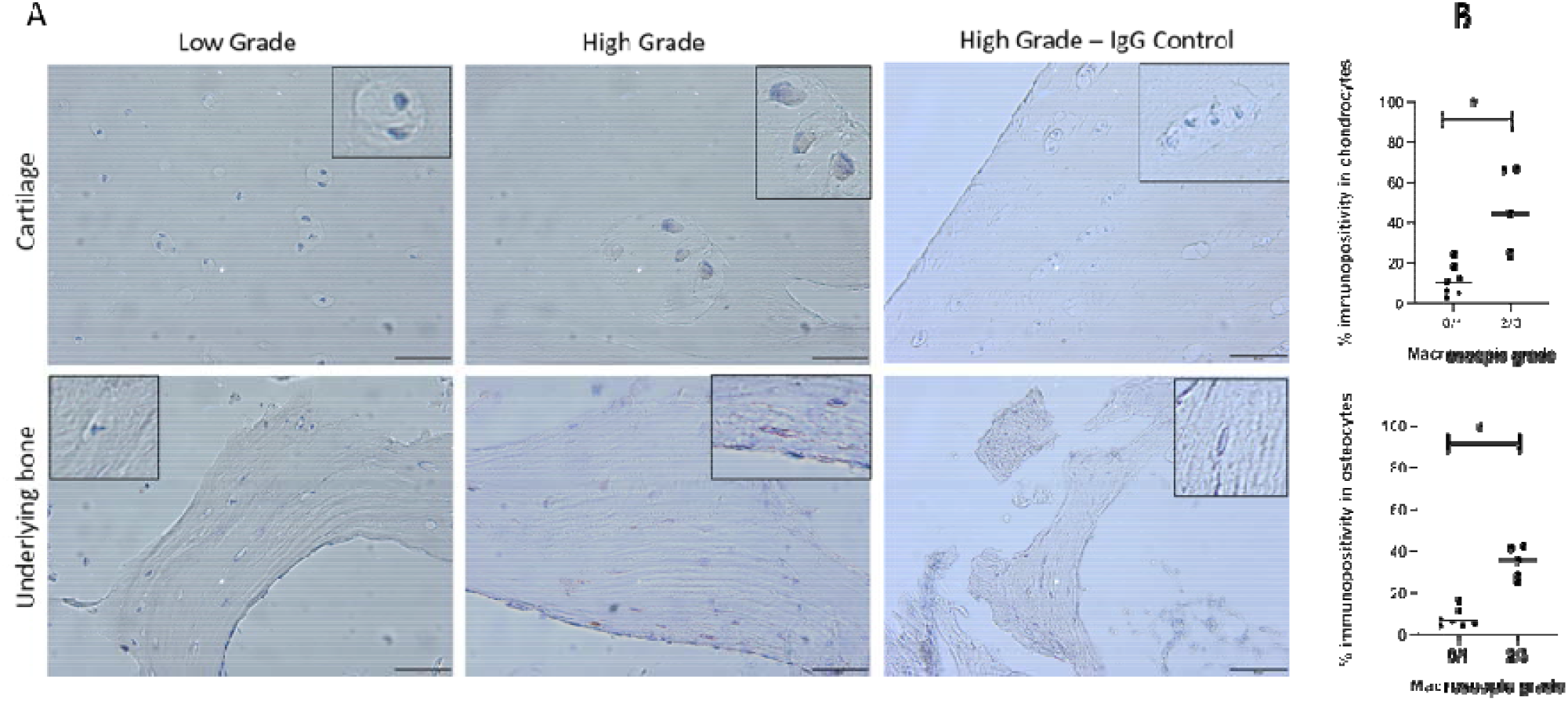
Immunohistochemistry localisation of TNFAIP8 within human osteochondral tissues. A: TNFAIP8 immunohistochemistry staining was identified in chondrocytes and osteocytes particularly in highly degraded (High Grade) tissues, IgG controls were negative, Middle zone cartilage shown within images. Scale bar = 50μm. (Insert shows zoomed cells) B: Percentage immunopositivity

#### Genetic correlations

The inter-study AA genetic correlation was reasonable (rg=0.57 [0.05-1.09] but the estimate was unreliable due to the small size of the RS GWAS (Supplementary Table 8). There was a moderate genetic correlation between AA and HOA (r_g_=0.26 [95% CI 0.10-0.43]) and minimum joint space width (r_g_= -0.31 [95% CI -0.46--0.15]) with an inverse relationship with the latter as expected. There was no or very limited evidence of a genetic correlation with hip pain, height, body mass index, bone mineral density or fracture (Supplementary Table 8).

### Mendelian randomisation

The 8 SNPs identified by LD clumping provided our genetic instrument for AA (for SNP effects in the outcome GWAS see Supplementary Table 3). The mean F-statistic was 31.5 indicating acceptable instrument strength. IVW analysis provided weak evidence of an effect of increasing AA on HOA risk (OR per SD change in AA 1.86 [1.09-3.15]) (Table 3). Sensitivity analyses, including MR Egger, weighted median, simple mode and weighted mode showed no evidence of a causal effect of AA on HOA risk (Figure 3). IVW and MR Egger Q statistics were 56.1 and 54.4 indicating heterogeneity and possible pleiotropy. Further sensitivity analyses, in the form of leave one out and single SNP analyses were used, in particular to assess the impact of rs561578905 which was identified by LD clumping but not by COJO, these revealed similar effect estimates (Supplementary Figures 8&9) and suggested no single SNP was responsible for the heterogeneity.

**Table 3.** Bi-directional Mendelian randomisation results comparing the causal effects between alpha angle and hip osteoarthritis.

| MR Method | Exposure AA, Outcome HOA |  | Exposure HOA, Outcome AA |  |
| --- | --- | --- | --- | --- |
|  | OR [95% CI] | *P* | Beta [95% CI] | *P* |
| Inverse variance weighted | 1.84 [1.14-2.96] | 0.01 | 0.09 [0.04-0.13] | 4.58 x 10<sup>-05</sup> |
| MR Egger | 1.22 [0.18-8.37]<sup>†</sup> | 0.84 | 0.15 [0.01-0.30]<sup>‡</sup> | 0.05 |
| Weighted median | 1.22 [0.93-1.59] | 0.16 | 0.08 [0.04-0.12] | 7.77 x 10<sup>-05</sup> |
| Simple mode | 1.33 [0.91-1.93] | 0.16 | 0.10 [0.00-0.19] | 0.05 |
| Weighted mode | 1.18 [0.92-1.51] | 0.27 | 0.12 [0.02-0.21] | 0.02 |

<sup>†</sup>MR Egger intercept 0.02, p-value 0.68. <sup>‡</sup>MR Egger intercept -0.01 p-value 0.34. AA – alpha angle, HOA – UK Biobank GWAS of hospital diagnosed hip osteoarthritis, MR – Mendelian randomisation. OR – odds ratio, per standard deviation change in alpha angle. Beta – per doubling in odds of hip osteoarthritis.

**Figure 3.**
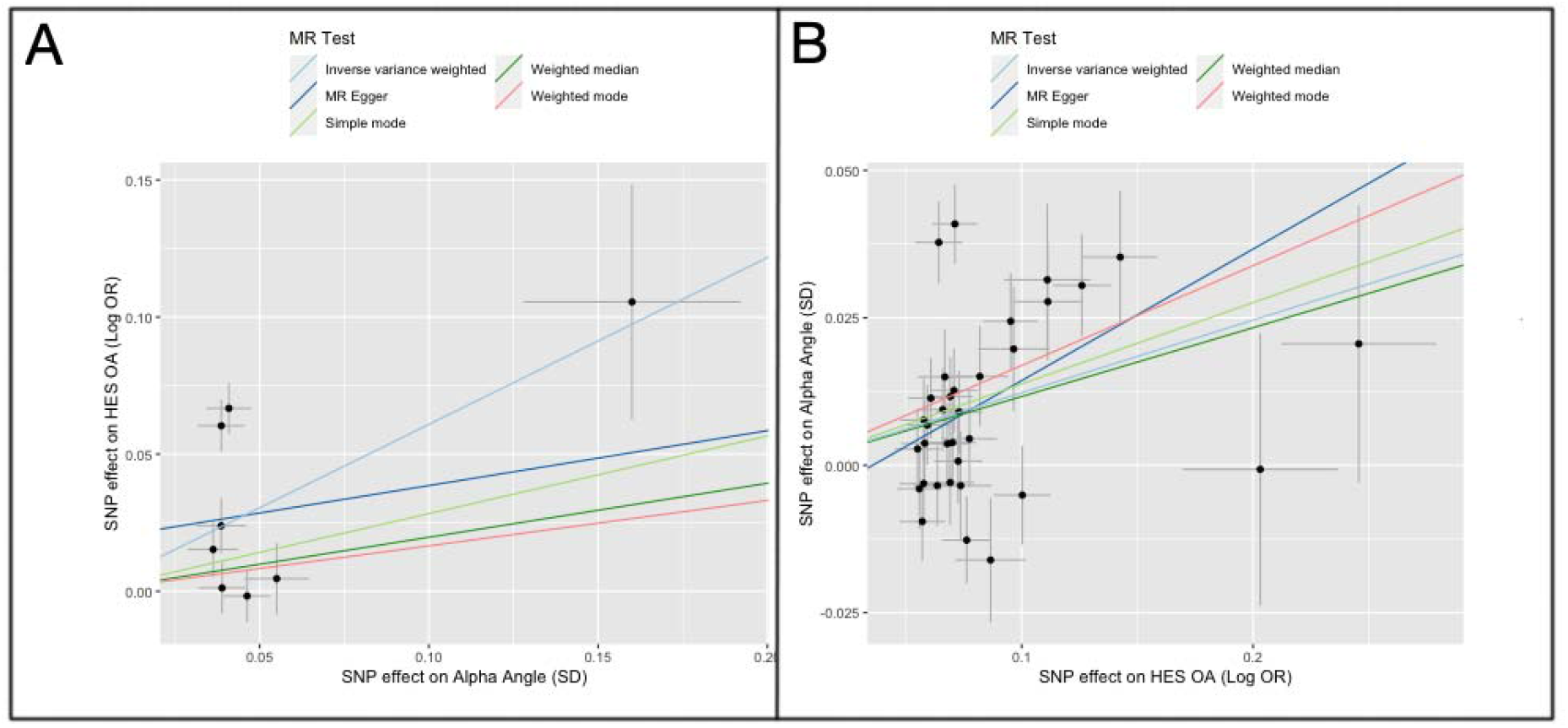
Bi-directional Mendelian randomisation results comparing the causal effects between alpha angle and hospital diagnosed hip osteoarthritis in the UK Biobank study. A – shows the MR analyses using eight genetic instruments for alpha angle (AA) as the exposure and hospital diagnosed hip osteoarthritis (HOA) as the outcome. B – shows the MR analyses using thirty-four genetic instruments for HOA as the exposure and AA as the outcome.

For MR analyses in the opposite direction, the HOA GWAS (323,948 participants) identified 34 independent SNPs following LD clumping (mean F-statistic 45.0) (Supplementary Table 9). In IVW analyses, genetic instruments for HOA showed a causal effect on AA (β 0.09 [0.04-0.13], β is SD change in AA per doubling in odds of OA) (Table 3). Sensitivity analyses were broadly in agreement (Figure 3). IVW and MR Egger Q statistics were 97.6 and 94.8 indicating heterogeneity.

MR-CAUSE analyses, which use whole GWAS summary statistics, were performed to examine for causal effects and correlated pleiotropy (33). For AA versus HOA there was only weak evidence the causal model (model 2) performed better than the null model (model 1) (expected log pointwise predictive density (ELPD) -3.80, p=0.07, an ELPD ≤;0 suggests model 2 fits the data better than model 1) (Supplementary Table 10). For HOA versus AA, there was stronger evidence that the causal model (model 2) performed better than the null (model 1) (ELPD -7.12, p=0.03), and better than the model assuming correlated pleiotropy (sharing model) (ELPD -3.65, p=0.02).

## Discussion

This is the first GWAS meta-analysis of AA, which identified eight loci and indicated a heritable component of 10%. *TGFA, TNFAIP8, CLDN20-RP11-477D19-TFB1M, LMX1B, GRK5, SOX5, CYP19A1* and *UQCC1* were implicated in increasing AA with *TNFAIP8* showing the strongest gene-SNP relationship. Despite strong evidence of observational associations, bi-directional two-sample MR analyses provided limited evidence of a causal association between increasing AA and the development of HOA, but rather showed greater evidence that a genetic predisposition to HOA causes an increase in AA, as measured by DXA in this subject cohort.

Of these eight loci, *TNFAIP8* colocalized with *cis*-eQTL expression in chondrocytes obtained from highly degraded, but not healthy, cartilage of the same individual. This suggestion that *TNFAIP8* is preferentially expressed in degraded cartilage was further explored by subsequent immunohistochemistry staining which showed greater expression of *TNFAIP8* in chondrocytes and osteocytes from degraded joint tissue. TNFAIP8 is a tumour necrosis factor binding protein forming part of inflammatory, catabolic and neuro-sensitisation pathways during the pathogenesis of OA, and is also involved in cell apoptosis (36). Inflammatory changes are well recognised in HOA (37), and our findings provide evidence to support the hypothesis that TNFAIP8 expression in osteocytes/chondrocytes contributes to hip shape remodelling which increases AA, leading to the appearance of cam morphology. How hip shape and an individual’s AA changes over time is not well understood and it could be that the observed shape variation arises in later life as part of the HOA process, as distinct to cam morphology caused by altered shape development in adolescence (9).

Rs62578126 and rs1048584 loci showed evidence of colocalisation with cis-eQTL signals for *LMX1B* and *CLDN20-RP11-477D19* -*TFB1M* in adipose tissue and fibroblasts respectively. *LMX1B* is the gene responsible for Nail-patella syndrome, which features poorly developed nails, patella and multiple limb malformations, and when knocked out in mice is associated with abnormal ventral limb development (38). *TFB1M* is important in preventing oxidative stress in mitochondria in the context of osteoarthritis (39). *CLDN20* is from the claudin family which are known to regulate osteoblast activity (40). *RP11-477D19* was also identified through co-localisation although little is known about its function.

The other SNPs provided no specific evidence of a causal gene through eQTL or colocalisation analyses. In these cases, we highlight that many of the closest genes identified have previously been implicated in limb development and OA. An intronic variant of *UQCC1* has been associated with developmental dysplasia of the hip (DDH) in a Han Chinese population (41). Interestingly, our *UQCC1* SNP (rs4911180) was in high LD with the lead *GDF5* OA SNP (r^2^ 0.79) from previous GWAS (2). The *UQCC1-GDF5* locus has been implicated in abnormal limb development and OA with both genes commonly expressed in chondrocytes (42-44). *TGFA* (rs7571789) is a growth factor which has been shown to be expressed in developing limbs in chicks (45) and important in the development of OA (46). *SOX5* (rs561578905) has previously been shown to be critical in joint morphogenesis through its action on growth plate and articular chondrocytes (47). *CYP19A1* (rs146939415) has been associated with large joint osteoarthritis and is thought to act via aromatase inhibition (48). Finally, *GRK5* is thought to regulate cartilage degradation and might be a possible therapeutic target for OA (49).

The eight independent AA SNPs had acceptable instrument strength when combined in subsequent MR analyses. However, there was only weak evidence of a causal link between increased AA and HOA. Interestingly, our bidirectional MR study provides stronger evidence that a genetic predisposition for HOA causes a higher AA, suggesting that the morphological features identified in this cohort may develop as part of, or in parallel to, the HOA process. Modelling changes are recognised by Croft grading in late-stage HOA (11), however we are not aware of any previous reports describing cam morphology as a specific feature of HOA. That said, in the same set of DXA images, we have recently found that hip shape changes suggestive of cam morphology are associated with more severe forms of HOA (50).

Although the evidence for a causal effect of a genetic predisposition to HOA on AA was somewhat stronger than that of AA on HOA, it should be noted that our genetic instrument for HOA was stronger than that for AA, reflecting the greater number of SNPs, so caution needs to be exercised in comparing these effects. Moreover, given that the age of our cohort was 40 to 69 years of age at inception, it could be that variation in AA largely reflected modelling changes that are part of the HOA disease process as opposed to cam morphology developing in early life. Alternatively, rather than bi-directional causal effects, it may be that our findings reflect common genetic pathways involved in the development of AA and HOA causing them to develop in parallel rather than as a consequence of one another. Though MR-CAUSE analysis favoured a causal over a shared model, this was only supported by weak evidence, and given the disparity in instrument strength between the two traits it is difficult to reach any firm conclusions. Nevertheless, to the extent that a causal effect of a genetic predisposition of HOA on AA and/or shared genetic pathways contribute to associations between cam morphology and HOA, our results suggest that HOA should not necessarily be attributed to cam morphology especially in individuals where they might co-exist. This has implications when considering hip shape augmenting surgery in an older adult population.

We report the first GWAS meta-analysis for AA in individuals from UKB and RS. Though we used continuous AA as a proxy measure for cam morphology, there are several limitations to this approach. For example, measuring AA on anterior-posterior images can be partially out of plane to the cam lesion, leading to an underestimation of size. However, our observational analyses suggest we are measuring a clinically relevant shape signal despite of this. When measuring AA in a population in later life there is the possibility that AA captures osteophytes or other features of OA. However, we rigorously excluded osteophytes and if these were included in our measures we might have expected to see a stronger causal relationship between AA and HOA. Though we used AA as a continuous measure to optimise statistical power, this method has less clinical relevance than dichotomising into the presence or absence of cam morphology based on a pre-defined cut-off (2, 21). Although, we found similar observational relationship between AA and cam morphology, and HOA outcomes irrespective of whether we used a continuous (AA) or binary (cam) measure. Moreover, sensitivity analyses based on a binary AA variable showed similar but underpowered GWAS results. Further work is needed to recruit hip imaging cohorts that are closer to UKB in terms of scale and phenotyping to allow for further replication of our results, and to extend our findings to more ethnically diverse populations. Finally, as with any MR study, several assumptions need to be made: the relevance assumption is satisfied by our ample F-statistics but the independence and exclusion restriction assumptions are harder to test (14). Several sensitivity analyses were performed to examine for possible pleiotropy which suggested this was present as has been discussed.

In conclusion, using a novel GWAS meta-analysis of AA, our study suggests that causal relationships between AA and HOA, and particularly a genetic predisposition for HOA and AA, contribute to observational associations between HOA and cam morphology. Changes in AA as a consequence of HOA development may involve up-regulation of inflammatory/catabolic pathways, given our observation that *TNFAIP8*, one of the top AA-associated loci, was preferentially expressed in degraded human articular cartilage and bone. Further studies are justified to explore the contribution of increased AA to clinical consequence of HOA, and to determine whether targeting the underlying molecular mechanisms might prove useful in ameliorating these.

## Supporting information

Supplementary Methods, Supplementary Figures 1-9, Strobe checklist

## Data Availability

The alpha angle GWAS meta-analysis summary statistics are to be uploaded to the GWAS catalogue (https://www.ebi.ac.uk/gwas/). The individual level data from this study will be available from UK Biobank in a forthcoming data release. Users must be registered with UK Biobank to access their resources (https://bbams.ndph.ox.ac.uk/ams/).

https://bbams.ndph.ox.ac.uk/ams/

## Acknowledgements

The authors would like to thank the Musculoskeletal Research Unit patient and public involvement group at the University of Bristol for their input into planning our research and Dr Martin Williams, Consultant Musculoskeletal Radiologist North Bristol NHS Trust, who provided substantial training and expertise for this study. This work has been conducted using the UK Biobank resource (application number 17295). The authors would like to thank the study participants, the staff from the Rotterdam Study and the participating general practitioners and pharmacists. The generation and management of GWAS genotype data for the Rotterdam Study (RSI, RSII, RSIII) was executed by the Human Genotyping Facility of the Genetic Laboratory of the Department of Internal Medicine, Erasmus MC, Rotterdam, The Netherlands. The authors would like to thank Pascal Arp, Mila Jhamai, Marijn Verkerk, Lizbeth Herrera and Marjolein Peters, MSc, and Carolina Medina-Gomez, MSc, for their help in creating the GWAS database, and Linda Broer PhD, for the creation of the imputed data. Mathijs Versteeg for creating the alpha angle data.

## Funding and grant award information

BGF is supported by a Medical Research Council (MRC) clinical research training fellowship (MR/S021280/1). RE, MF, FS are supported, and this work is funded by a Wellcome Trust collaborative award (reference number 209233). CL is funded by a Sir Henry Dale Fellowship jointly funded by the Wellcome Trust and the Royal Society (223267/Z/21/Z). This research was funded in whole, or in part, by the Wellcome Trust [Grant numbers 080280/Z/06/Z, 20378/Z/16/Z, 223267/Z/21/Z]. For the purpose of open access, the authors have applied a CC BY public copyright licence to any Author Accepted Manuscript version arising from this submission. NCH acknowledges support from the MRC and NIHR Southampton Biomedical Research Centre, University of Southampton and University Hospital Southampton. BGF, MF, AEH, GDS, JHT work in the MRC Integrative Epidemiology Unit at the University of Bristol, which is supported by the MRC (MC_UU_00011/1). JPK is funded by a National Health and Medical Research Council (Australia) Investigator grant (GNT1177938).

## Ethical approval statement

The National Information Governance Board for Health and Social Care and Northwest Multi-Centre Research Ethics Committee (11/NW/0382) and UK Biobank Ethics Advisory committee gave ethical approval for all work in this study undertaken with UK Biobank data (UK Biobank application number 17295). The South Yorkshire and North Derbyshire Musculoskeletal Biobank (REC reference: 20/SC/0144) ethics committee gave ethical approval for the immunohistochemistry experiments in this study. The Medical Ethics Committee of the Erasmus MC (registration number MEC 02.1015) and the Dutch Ministry of Health, Welfare and Sport (Population Screening Act WBO, license number 1071272-159521-PG) gave ethical approval for all work undertaken with data from the Rotterdam Study included in this study. The Rotterdam Study has been entered into the Netherlands National Trial Register (NTR; www.trialregister.nl) and into the WHO International Clinical Trials Registry Platform (ICTRP; www.who.int/ictrp/network/primary/en/) under shared catalogue number NTR6831. All participants provided informed consent for this study.

## References

1. Faber BG, Frysz M, Tobias JH. Unpicking observational relationships between hip shape and osteoarthritis: hype or hope? Curr Opin Rheumatol. 2020;32(1):110–8.

2. Agricola R, Heijboer MP, Bierma-Zeinstra SMA, Verhaar JAN, Weinans H, Waarsing JH. Cam impingement causes osteoarthritis of the hip: a nationwide prospective cohort study (CHECK). Annals of the Rheumatic Diseases. 2013;72(6):918–23.

3. Thomas GE, Palmer AJ, Batra RN, Kiran A, Hart D, Spector T, et al. Subclinical deformities of the hip are significant predictors of radiographic osteoarthritis and joint replacement in women. A 20 year longitudinal cohort study. Osteoarthritis Cartilage. 2014;22(10):1504–10.

4. Griffin DR, Dickenson EJ, Wall PDH, Achana F, Donovan JL, Griffin J, et al. Hip arthroscopy versus best conservative care for the treatment of femoroacetabular impingement syndrome (UK FASHIoN): a multicentre randomised controlled trial. Lancet. 2018;391(10136):2225–35.

5. Palmer AJR, Ayyar Gupta V, Fernquest S, Rombach I, Dutton SJ, Mansour R, et al. Arthroscopic hip surgery compared with physiotherapy and activity modification for the treatment of symptomatic femoroacetabular impingement: multicentre randomised controlled trial. BMJ. 2019;364:l185.

6. Jepsen P, Johnsen SP, Gillman MW, Sorensen HT. Interpretation of observational studies. Heart. 2004;90(8):956–60.

7. Lee H, Aronson J. Association or causation? How do we ever know? [Available from: https://catalogofbias.org/2019/03/05/association-or-causation-how-do-we-ever-know/.

8. Notzli HP, Wyss TF, Stoecklin CH, Schmid MR, Treiber K, Hodler J. The contour of the femoral head-neck junction as a predictor for the risk of anterior impingement. J Bone Joint Surg Br. 2002;84(4):556–60.

9. van Klij P, Heijboer MP, Ginai AZ, Verhaar JAN, Waarsing JH, Agricola R. Cam morphology in young male football players mostly develops before proximal femoral growth plate closure: a prospective study with 5-yearfollow-up. Br J Sports Med. 2019;53(9):532–8.

10. Monazzam S, Bomar JD, Pennock AT. Idiopathic Cam Morphology Is Not Caused by Subclinical Slipped Capital Femoral Epiphysis: An MRI and CT Study. Orthop J Sports Med. 2013;1(7):2325967113512467.

11. Croft P, Cooper C, Wickham C, Coggon D. Defining osteoarthritis of the hip for epidemiologic studies. American Journal of Epidemiology. 1990;132(3):514–22.

12. Ganz R, Parvizi J, Beck M, Leunig M, Nötzli H, Siebenrock KA. Femoroacetabular Impingement: A Cause for Osteoarthritis of the Hip. Clinical Orthopaedics and Related Research. 2003;417:112–20.

13. Griffin DR, Dickenson EJ, O’Donnell J, Agricola R, Awan T, Beck M, et al. The Warwick Agreement on femoroacetabular impingement syndrome (FAI syndrome): an international consensus statement. Br J Sports Med. 2016;50(19):1169–76.

14. Davies NM, Holmes MV, Davey Smith G. Reading Mendelian randomisation studies: a guide, glossary, and checklist for clinicians. BMJ. 2018;362:k601.

15. Littlejohns TJ, Holliday J, Gibson LM, Garratt S, Oesingmann N, Alfaro-Almagro F, et al. The UK Biobank imaging enhancement of 100,000 participants:[rationale, data collection, management and future directions. Nature Communications. 2020;11(1):2624.

16. Sanderson E, Glymour MM, Holmes MV, Kang H, Morrison J, Munafò MR, et al. Mendelian randomization. Nature Reviews Methods Primers. 2022;2(1):6.

17. Faber BG, Ebsim R, Saunders FR, Frysz M, Lindner C, Gregory JS, et al. A novel semi-automated classifier of hip osteoarthritis on DXA images shows expected relationships with clinical outcomes in UK Biobank. Rheumatology (Oxford). 2021.

18. Faber BG, Ebsim R, Saunders FR, Frysz M, Lindner C, Gregory JS, et al. Osteophyte size and location on hip DXA scans are associated with hip pain: findings from a cross sectional study in UK Biobank. Bone. 2021:116146.

19. Faber BG, Ebsim R, Saunders FR, Frysz M, Davey Smith G, Cootes T, et al. Deriving alpha angle from anterior-posterior dual-energy x-ray absorptiometry scans: an automated and validated approach. Wellcome Open Research. 2021 (https://wellcomeopenresearch.org/articles/6-60/v1).

20. Faber BG, Ebsim R, Saunders FR, Frysz M, Gregory JS, Aspden RM, et al. Cam morphology but neither acetabular dysplasia nor pincer morphology is associated with osteophytosis throughout the hip: findings from a cross-sectional study in UK Biobank. Osteoarthritis Cartilage. 2021;29(11):1521–9.

21. Saberi Hosnijeh F, Zuiderwijk ME, Versteeg M, Smeele HT, Hofman A, Uitterlinden AG, et al. Cam Deformity and Acetabular Dysplasia as Risk Factors for Hip Osteoarthritis. Arthritis Rheumatol. 2017;69(1):86–93.

22. Rangamaran VR, Uppili B, Gopal D, Ramalingam K. EasyQC: Tool with Interactive User Interface for Efficient Next-Generation Sequencing Data Quality Control. J Comput Biol. 2018;25(12):1301–11.

23. Willer CJ, Li Y, Abecasis GR. METAL: fast and efficient meta-analysis of genomewide association scans. Bioinformatics. 2010;26(17):2190–1.

24. Yang J, Ferreira T, Morris AP, Medland SE, Genetic Investigation of ATC, Replication DIG, et al. Conditional and joint multiple-SNP analysis of GWAS summary statistics identifies additional variants influencing complex traits. Nat Genet. 2012;44(4):369-75, S1-3.

25. Bulik-Sullivan B, Finucane HK, Anttila V, Gusev A, Day FR, Loh PR, et al. An atlas of genetic correlations across human diseases and traits. Nat Genet. 2015;47(11):1236–41.

26. Consortium GT, Laboratory DA, Coordinating Center -Analysis Working G, Statistical Methods groups-Analysis Working G, Enhancing Gg, Fund NIHC, et al. Genetic effects on gene expression across human tissues. Nature. 2017;550(7675):204–13.

27. Panjwani N, Wang F, Mastromatteo S, Bao A, Wang C, He G, et al. LocusFocus: Web-based colocalization for the annotation and functional follow-up of GWAS. PLoS Comput Biol. 2020;16(10):e1008336.

28. Giambartolomei C, Vukcevic D, Schadt EE, Franke L, Hingorani AD, Wallace C, et al. Bayesian test for colocalisation between pairs of genetic association studies using summary statistics. PLoS Genet. 2014;10(5):e1004383.

29. Steinberg J, Southam L, Roumeliotis TI, Clark MJ, Jayasuriya RL, Swift D, et al. A molecular quantitative trait locus map for osteoarthritis. Nat Commun. 2021;12(1):1309.

30. Boyle AP, Hong EL, Hariharan M, Cheng Y, Schaub MA, Kasowski M, et al. Annotation of functional variation in personal genomes using RegulomeDB. Genome Res. 2012;22(9):1790–7.

31. Skrivankova VW, Richmond RC, Woolf BAR, Yarmolinsky J, Davies NM, Swanson SA, et al. Strengthening the Reporting of Observational Studies in Epidemiology Using Mendelian Randomization: The STROBE-MR Statement. JAMA. 2021;326(16):1614–21.

32. Hemani G, Tilling K, Davey Smith G. Orienting the causal relationship between imprecisely measured traits using GWAS summary data. PLoS Genet. 2017;13(11):e1007081.

33. Morrison J, Knoblauch N, Marcus JH, Stephens M, He X. Mendelian randomization accounting for correlated and uncorrelated pleiotropic effects using genome-wide summary statistics. Nat Genet. 2020;52(7):740–7.

34. Agricola R, Waarsing JH, Thomas GE, Carr AJ, Reijman M, Bierma-Zeinstra SM, et al. Cam impingement: defining the presence of a cam deformity by the alpha angle: data from the CHECK cohort and Chingford cohort. Osteoarthritis Cartilage. 2014;22(2):218–25.

35. Boer CG, Hatzikotoulas K, Southam L, Stefansdottir L, Zhang Y, Coutinho de Almeida R, et al. Deciphering osteoarthritis genetics across 826,690 individuals from 9 populations. Cell. 2021;184(18):4784–818 e17.

36. Zhang L, Liu R, Luan YY, Yao YM. Tumor Necrosis Factor-alpha Induced Protein 8: Pathophysiology, Clinical Significance, and Regulatory Mechanism. Int J Biol Sci. 2018;14(4):398–405.

37. Hunter DJ, Bierma-Zeinstra S. Osteoarthritis. Lancet. 2019;393(10182):1745–59.

38. Feenstra JM, Kanaya K, Pira CU, Hoffman SE, Eppey RJ, Oberg KC. Detection of genes regulated by Lmx1b during limb dorsalization. Dev Growth Differ. 2012;54(4):451–62.

39. He Y, Wu Z, Xu L, Xu K, Chen Z, Ran J, et al. The role of SIRT3-mediated mitochondrial homeostasis in osteoarthritis. Cell Mol Life Sci. 2020;77(19):3729–43.

40. Lindsey RC, Xing W, Pourteymoor S, Godwin C, Gow A, Mohan S. Novel Role for Claudin-11 in the Regulation of Osteoblasts via Modulation of ADAM10-Mediated Notch Signaling. J Bone Miner Res. 2019;34(10):1910–22.

41. Sun Y, Wang C, Hao Z, Dai J, Chen D, Xu Z, et al. A common variant of ubiquinol-cytochrome c reductase complex is associated with DDH. PLoS One. 2015;10(4):e0120212.

42. Harsanyi S, Zamborsky R, Kokavec M, Danisovic L. Genetics of developmental dysplasia of the hip. European Journal of Medical Genetics. 2020;63(9):103990.

43. Sun K, Guo J, Yao X, Guo Z, Guo F. Growth differentiation factor 5 in cartilage and osteoarthritis: A possible therapeutic candidate. Cell Prolif. 2021;54(3):e12998.

44. Hatzikotoulas K, Roposch A, Consortium DDHCC, Shah KM, Clark MJ, Bratherton S, et al. Genome-wide association study of developmental dysplasia of the hip identifies an association with GDF5. Commun Biol. 2018;1:56.

45. Dealy CN, Scranton V, Cheng HC. Roles of transforming growth factor-alpha and epidermal growth factor in chick limb development. Dev Biol. 1998;202(1):43–55.

46. Usmani SE, Ulici V, Pest MA, Hill TL, Welch ID, Beier F. Context-specific protection of TGFalpha null mice from osteoarthritis. Sci Rep. 2016;6:30434.

47. Dy P, Smits P, Silvester A, Penzo-Méndez A, Dumitriu B, Han Y, et al. Synovial joint morphogenesis requires the chondrogenic action of Sox5 and Sox6 in growth plate and articular cartilage. Developmental Biology. 2010;341(2):346–59.

48. Riancho JA, Garcia-Ibarbia C, Gravani A, Raine EV, Rodriguez-Fontenla C, Soto-Hermida A, et al. Common variations in estrogen-related genes are associated with severe large-joint osteoarthritis: a multicenter genetic and functional study. Osteoarthritis Cartilage. 2010;18(7):927–33.

49. Sueishi T, Akasaki Y, Goto N, Kurakazu I, Toya M, Kuwahara M, et al. GRK5 Inhibition Attenuates Cartilage Degradation via Decreased NF-kappaB Signaling. Arthritis Rheumatol. 2020;72(4):620–31.

50. Frysz M, Faber BG, Ebsim R, Saunders FR, Lindner C, Gregory JS, et al. Machine-learning derived acetabular dysplasia and cam morphology are features of severe hip osteoarthritis: findings from UK Biobank. J Bone Miner Res. 2022.

