## Supplementary Methods, Supplementary Figures 1-9, Strobe checklist for "Investigation of the genetic architecture of cam morphology, and its relationship with hip osteoarthritis, using alpha angle as a proxy measure": Supplementary materials 15.7.22.docx

STROBE Statement—Checklist of items that should be included in reports of ***cohort studies***

|  | Item No | Recommendation | Page No |
| --- | --- | --- | --- |
| **Title and abstract** | 1 | (*a*) Indicate the study’s design with a commonly used term in the title or the abstract | 1 |
|  |  | (*b*) Provide in the abstract an informative and balanced summary of what was done and what was found | 2 |
| Introduction | | | |
| Background/rationale | 2 | Explain the scientific background and rationale for the investigation being reported | 4 |
| Objectives | 3 | State specific objectives, including any prespecified hypotheses | 5 |
| Methods | | | |
| Study design | 4 | Present key elements of study design early in the paper | 6 |
| Setting | 5 | Describe the setting, locations, and relevant dates, including periods of recruitment, exposure, follow-up, and data collection | Supp Methods |
| Participants | 6 | (*a*) Give the eligibility criteria, and the sources and methods of selection of participants. Describe methods of follow-up | Supp Methods |
|  |  | (*b*) For matched studies, give matching criteria and number of exposed and unexposed | Supp Methods |
| Variables | 7 | Clearly define all outcomes, exposures, predictors, potential confounders, and effect modifiers. Give diagnostic criteria, if applicable | 6 |
| Data sources/ measurement | 8* | For each variable of interest, give sources of data and details of methods of assessment (measurement). Describe comparability of assessment methods if there is more than one group | 6-8 |
| Bias | 9 | Describe any efforts to address potential sources of bias | 7-8 |
| Study size | 10 | Explain how the study size was arrived at | 6 |
| Quantitative variables | 11 | Explain how quantitative variables were handled in the analyses. If applicable, describe which groupings were chosen and why | 6-8 |
| Statistical methods | 12 | (*a*) Describe all statistical methods, including those used to control for confounding | 6-8  & |
|  |  | (*b*) Describe any methods used to examine subgroups and interactions | Supp |
|  |  | (*c*) Explain how missing data were addressed | Methods |
|  |  | (*d*) If applicable, explain how loss to follow-up was addressed |  |
|  |  | (*e*) Describe any sensitivity analyses |  |
| Results | | |  |
| Participants | 13* | (a) Report numbers of individuals at each stage of study—eg numbers potentially eligible, examined for eligibility, confirmed eligible, included in the study, completing follow-up, and analysed | 9 |
|  |  | (b) Give reasons for non-participation at each stage | Supp |
|  |  | (c) Consider use of a flow diagram | Results |
| Descriptive data | 14* | (a) Give characteristics of study participants (eg demographic, clinical, social) and information on exposures and potential confounders | Supp |
|  |  | (b) Indicate number of participants with missing data for each variable of interest | Results |
|  |  | (c) Summarise follow-up time (eg, average and total amount) | Supp Table 1 |
| Outcome data | 15* | Report numbers of outcome events or summary measures over time | Supp Table 1 |

| Main results | 16 | (*a*) Give unadjusted estimates and, if applicable, confounder-adjusted estimates and their precision (eg, 95% confidence interval). Make clear which confounders were adjusted for and why they were included | Tab 1,2,3 |
| --- | --- | --- | --- |
|  |  | (*b*) Report category boundaries when continuous variables were categorized | 9-12 |
|  |  | (*c*) If relevant, consider translating estimates of relative risk into absolute risk for a meaningful time period |  |
| Other analyses | 17 | Report other analyses done—eg analyses of subgroups and interactions, and sensitivity analyses | 9-12 |
| Discussion | | | |
| Key results | 18 | Summarise key results with reference to study objectives | 13 |
| Limitations | 19 | Discuss limitations of the study, taking into account sources of potential bias or imprecision. Discuss both direction and magnitude of any potential bias | 15-16 |
| Interpretation | 20 | Give a cautious overall interpretation of results considering objectives, limitations, multiplicity of analyses, results from similar studies, and other relevant evidence | 13-17 |
| Generalisability | 21 | Discuss the generalisability (external validity) of the study results | 15-17 |
| Other information | | | |
| Funding | 22 | Give the source of funding and the role of the funders for the present study and, if applicable, for the original study on which the present article is based | 18-19 |

*Give information separately for exposed and unexposed groups.

**Note:** An Explanation and Elaboration article discusses each checklist item and gives methodological background and published examples of transparent reporting. The STROBE checklist is best used in conjunction with this article (freely available on the Web sites of PLoS Medicine at http://www.plosmedicine.org/, Annals of Internal Medicine at http://www.annals.org/, and Epidemiology at http://www.epidem.com/). Information on the STROBE Initiative is available at http://www.strobe-statement.org.

Supplementary methods:

UK Biobank

UKB recruited 500,000 adults prospectively between 2006-2010 who have undergone comprehensive genetic and physical phenotyping (http://biobank.ctsu.ox.ac.uk/crystal/) (1). The UKB extended imaging study has conducted hip DXA scans on ~50,000 individuals to date. Demographic information was obtained on the same day as the DXA scans.

*Outline points, radiographic hip osteoarthritis and alpha angle*

This study included those UKB participants with a left hip DXA scan (iDXA GE-Lunar, Madison, WI) available in December 2021 (2, 3). Custom software placed 85 points around the femoral head and acetabulum (Figure 1) (4, 5). All images were checked manually but only 10% of images required correction (6). At the same time osteophytes were annotated manually and excluded from the outline points, from which osteophyte grade was derived (7). JSN narrowing was calculated from minimum joint space width measured automatically between the outline points and combined with osteophyte grade to give an overall grade of radiographic HOA (rHOA) 0-4 (6). AA was calculated automatically using a publicly available and validated Python 3.0 script which utilises the aforementioned outline points (Figure 1) (8-10). Previous studies have shown good repeatability of these methods (concordance correlation coefficient 0.84) (10). When considering cam morphology as a binary outcome for sensitivity analyses this was defined as an AA ≥60°.

*Hospital diagnosed osteoarthritis and total hip replacement*

Hospital diagnosed HOA was derived from international classification of diseases (ICD) -9 &10 codes released in hospital episode statistics (HES) linked to UKB in January 2021. The full list of codes included in this study has previously been published (11). This outcome is referred to as HES OA. HES data has been collected since 1981 in Scotland, 1997 in England and 1998 in Wales. Total hip replacement (THR) data were derived from Office of Population Censuses and Survey (OPCS) -3&4 codes which are used to record operation procedures in HES. This study included the following OPCS-3 codes: 811, 810 and OPCS-4 codes: W371, W81, W391. This data was collected over the same period as described for HES OA. Both variables were binary and were not side specific.

Observational analyses

Demographic data are shown as mean and range for continuous variables and counts, and frequency for binary variables. Logistic regression was used to examine associations between AA, and rHOA (grade ≥2) and hip pain and HES OA, results are given as odds ratios (OR) with 95% confidence intervals (CI). Cox proportional hazard modelling was used to examine associations between AA and THR, results are given as hazard ratios (HR) with 95% CI. Covariates for the adjusted model included age, height, weight and sex. Statistical analysis used Stata version 16 (StataCorp, College Station, TX, USA).

Rotterdam Studies and their alpha angle measurements

The Rotterdam Study (RS) is a prospective population based cohort consisting of elderly inhabitants, 45 years and older, of the Omnoord district in the city of Rotterdam, the Netherlands. The design of the Rotterdam Study, data collection and genotyping has been described in detail in (12). Participants from the Rotterdam Study I (n=2,970), II (n=1,254) & III (n=1,254) were included in this study. Briefly, these are three phases of recruitment (I – 1989, those aged >55 years old, II – 2000, those aged >55 years old & III – 2006, those aged >45 years old).

Weight-bearing anteroposterior pelvic radiographs were obtained at 70 kV, a focus of 1,8, and a focus-to-film distance of 120 cm, using High resolution G 35 x 43-cm film (Fuji Photo Film Company, Kanagawa, Japan). Both of the participant’s feet were positioned in 10° internal rotation. The X-ray beam was centered on the umbilicus. All radiographs of the pelvis are digitized to DCM files. Before measuring the angles to detect cam impingement, the baseline x-rays were converted from DCM to JPEG files and have been edited with a colour filter. The radiographs were grouped per participant and read by pairs in chronological order, the order being known by the reader.

The shape of the proximal femur and of a part of the acetabulum was outlined using statistical shape modelling (SSM) software (ASM tool kit, Manchester University, Manchester, UK). A set of 23 points was positioned on anatomical landmarks along the surface of the bone on the anteroposterior radiographs. From this point set of the SSM software, the alpha angle was automatically calculated using Matlab (13). In brief, a fitting circle was drawn around the femoral head as precisely as possible. Second, a line was drawn through the centre of the femoral neck and head. Finally, a second line needs to be drawn, from the centre of the femoral head to the point where the superior surface of the femur head/neck departs from the circle. The alpha angle is the angle between the two drawn lines. The point sets were placed by four investigators (2 investigators placed the point set of RS1, 1 placed them for RS2 and 1 placed them for RS3). The ICC score for inter-observer reliability of the alpha angle was 0.50 (95% CI 0.26 – 0.68) and the ICC score for intra-observer reliability of the alpha angle was 0.91 (95% CI 0.86 – 0.94)

GWAS in UK Biobank

Genotyping, imputation and quality control (QC) were performed by UKB as previously described (14). Samples were genotyped using two genotyping arrays; Applied Biosystems UK BiLEVE Axiom Array by Affymetrix (49,950 participants) and Applied Biosystems UK Biobank Axiom Array (438,427 participants). Data were imputed using the HRC reference panel, and the merged UK10K and 1000 Genomes phase 3 reference panels in IMPUTE4.

A subset of Europeans individuals from UKB (1) was used for GWAS. Ancestry assignment of UK-Biobank participants was performed as follows: The UKB sample was projected onto the first 20 principal components estimated from the 1000 Genomes Phase 3 (1000G) project (where ancestry was known) using GCTA version 1.93.2. Projections used a curated set of 38,512 LD-pruned HapMap 3 Release 3 (HM3)REF bi-allelic SNPs that were shared between the 1000G and UKB genotyped datasets (i.e. MAF > 1%, minor allele count > 5, genotyping call rate > 95%, Hardy-Weinberg P > 1x10^-6^, and regions of extensive LD removed). Uniform Manifold Approximation and Projection for Dimension Reduction (UMAP) was used in conjunction with the first 20 principal components to cluster 486,445 individuals using the following parameters: min_dist=0.0001, n_components=3, n_neighbors=45, random_state=10293082. UKB participants that clustered together with the 1000G European sub-populations were manually identified by visual inspection (N=461,920) and used for downstream genetic analyses.

We tested associations between genetic variants and standardised (mean=1, SD=1) AA and HES OA assuming an additive allelic effect using linear mixed model (LMM) implemented in BOLT-LMM (v2.3) to account for cryptic population structure and relatedness, correcting each trait for age, sex, genotyping array and ancestry informative principal components 1 - 20 as previously described. GWAS involved high quality genome-wide imputed v3 genetic data (i.e. ~12 million SNPs, INFO > 0.3, MAF > 0.01%) measured in 38,173 related Europeans (for AA and cam morphology defined as AA ≥60°) and 323,948 (for HES OA) from the UK-Biobank Study. No individuals were in both GWAS as there was no sample overlap. Independent SNPs were annotated to the closest gene using the hg19 gene range list in dbsnp (<https://www.ncbi.nlm.nih.gov/SNP/>).

Locus zoom plots from the AA meta-analysis were generated and used to visually inspect the patterns of association at each locus.

GWAS in Rotterdam

Genotypes from RS participates were imputed to the Haplotype Reference Consortium reference panel (V.1.0) using the Michigan Imputation Server (15). We assessed genetic associations in each RS sub-cohort (RSI, RSII and RSIII) using linear regression models of the standardised alpha angle residuals (adjusted for age and sex) and additionally adjusted for the first four genetic principal components. RVtests (16) was used for the GWAS analyses and results were quality controlled using EasyQC (17). Variants with an imputation quality <0.3, minor allele frequency <0.05 or effective allele count <5 were excluded and genomic control correction was applied to all SE and p values.

Genetic correlation and heritability

LD score regression (LDSR) was used to estimate the genetic correlation between AA in UKB and RS. In addition, heritability and further genetic correlations were estimated for the AA meta-analysis GWAS and HOA, THR (18), minimum joint space width (mJSW) (two GWAS used: (i) mJSW meta-analysis between UKB, MrOS, SOF, RS, and (ii) mJSW meta-analysis between MrOS, SOF, RS because UKB mJSW was derived on DXA as opposed to radiographs), acute and chronic hip pain (UKB only GWAS), fracture (19), height (20), BMI (21), bone mineral density (BMD) (based on lumbar spine and femoral neck BMD on DXA in UKB) and estimated bone mineral density (eBMD) (22). We used well established methods (23) facilitated by pre-computed LD scores from European populations ([https://data.broadinstitute.org/alkesgroup/LDSCORE /eur_w_ld_chr.tar.bz2](https://data.broadinstitute.org/alkesgroup/LDSCORE%20/eur_w_ld_chr.tar.bz2)). Summary statistics were munged with their package munge.py (therefore excluding major histocompatibility regions) prior to LDSR.

Immunohistochemistry

To investigate expression of TNFAIP8 within human osteoarthritic joints and investigate potential differential expression between non-degenerate and degenerate regions, immunohistochemistry was utilised to localise TNFAIP8 expression in patient osteochondral tissue. Osteochondral tissue samples were harvested from the tibial plateau or distal femoral condyle from 4 patients with knee OA (Ethics approval South Yorkshire and North Derbyshire Musculoskeletal Biobank (REC reference: 20/SC/0144)) 1 male, 3 female, age 59-78. Tissue was macroscopically graded according to the International Cartilage Repair Society grading immediately following harvest and tissue region paired samples of less (0/1) and highly (2/3) degraded osteochondral tissue plugs were harvested, formalin fixed, EDTA decalcified and paraffin embedded (24, 25). Four-micron sections were probed using immunohistochemistry for TNFAIP8 expression. Immunohistochemistry was performed using standard methodology as previously published (26), utilising chymotrypsin enzyme retrieval, 2hr 25% v/v Rabbit serum in 1% w/v BSA in TBS, overnight incubation in 1:100 dilution of mouse monoclonal TNFAIP8 (Invitrogen, MA5-27345) together with equivalent IgG concentration controls. Following overnight incubation and washes in tris buffered Saline (TBS), 30 mins 1:400 biotinylated rabbit anti-mouse (Abcam) secondary antibody was performed. Washed in TBS and signal amplified with ABC (Vector) and detected using DAB (Sigma), Images were analysed on Olympus BX60 microscope and images captured using CellSens software (Olympus). Percentage immunopositivity was calculated for articular chondrocytes and osteocytes within the underlying bone by counting at least 200 cells in each region, and Mann Whitney U test performed using Graph Pad Prism v8.1.1 to determine the statistical evidence of association between low- and high-grade tissues.

Supplementary Figures:

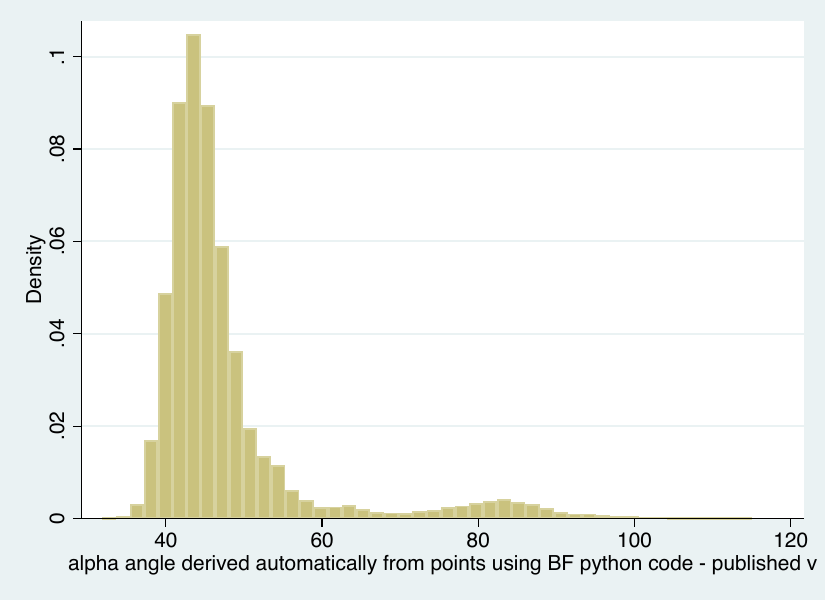

Supplementary Figure 1: Alpha angle distribution in UK Biobank

DXA and AA measure available in UKB (n=40,337). Observational associations done in this population

Those that have an AA measure and pas genetic quality control in UK Biobank

(n = 38,173)

Alpha Angle GWAS meta-analysis (n=44,214)

Individuals who fail genetic quality control in UK Biobank (n=2,164)

Those that have an AA measure and pass genetic quality control in the Rotterdam Studies (n = 6,041)

Supplementary Figure 2. A flow chart of the populations used in this study.

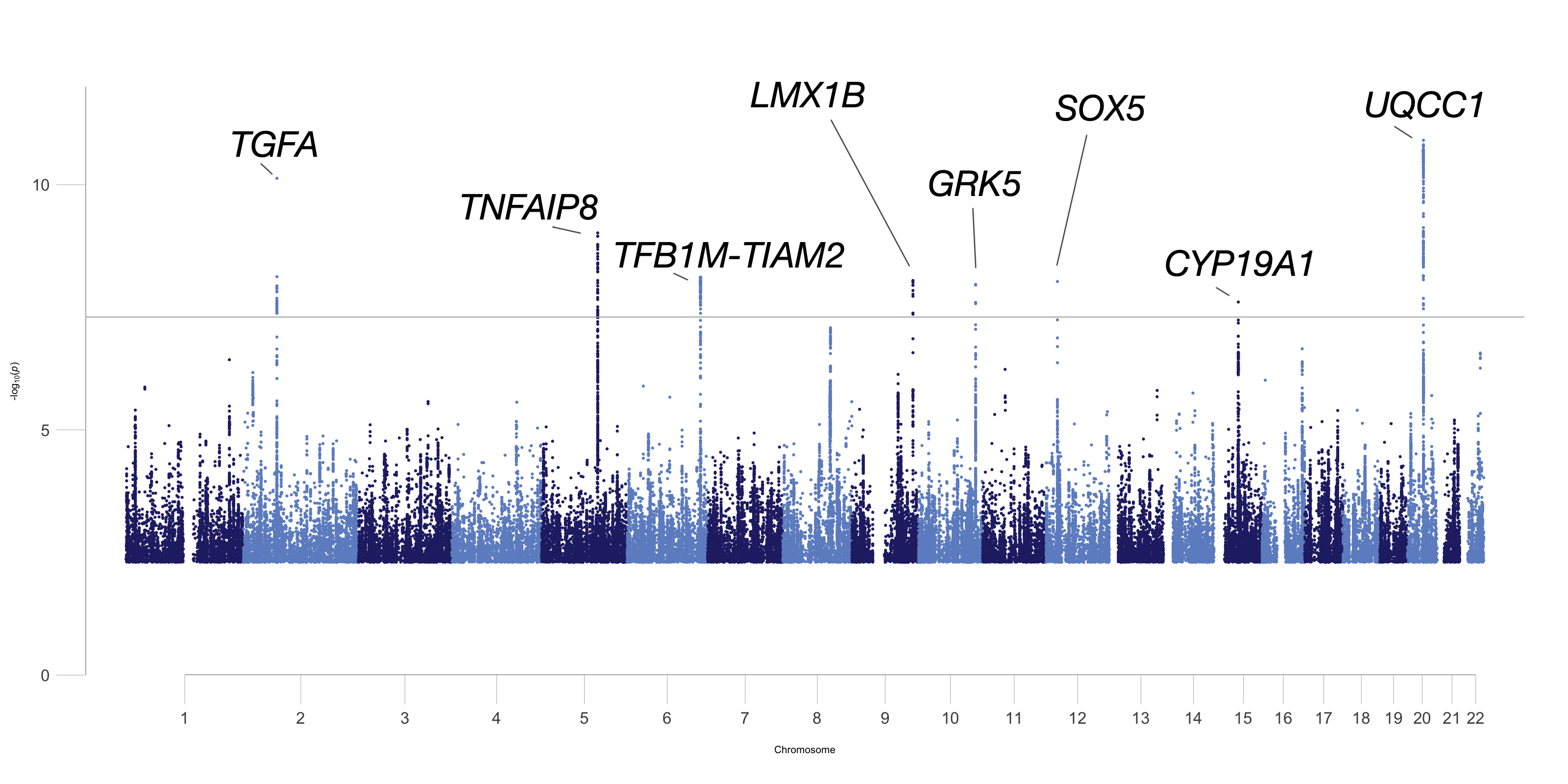

Supplementary Figure 3. A Manhattan Plot describing the alpha angle GWAS meta-analysis. The closest genes label the independent genetic loci that meet genome-wide significance.

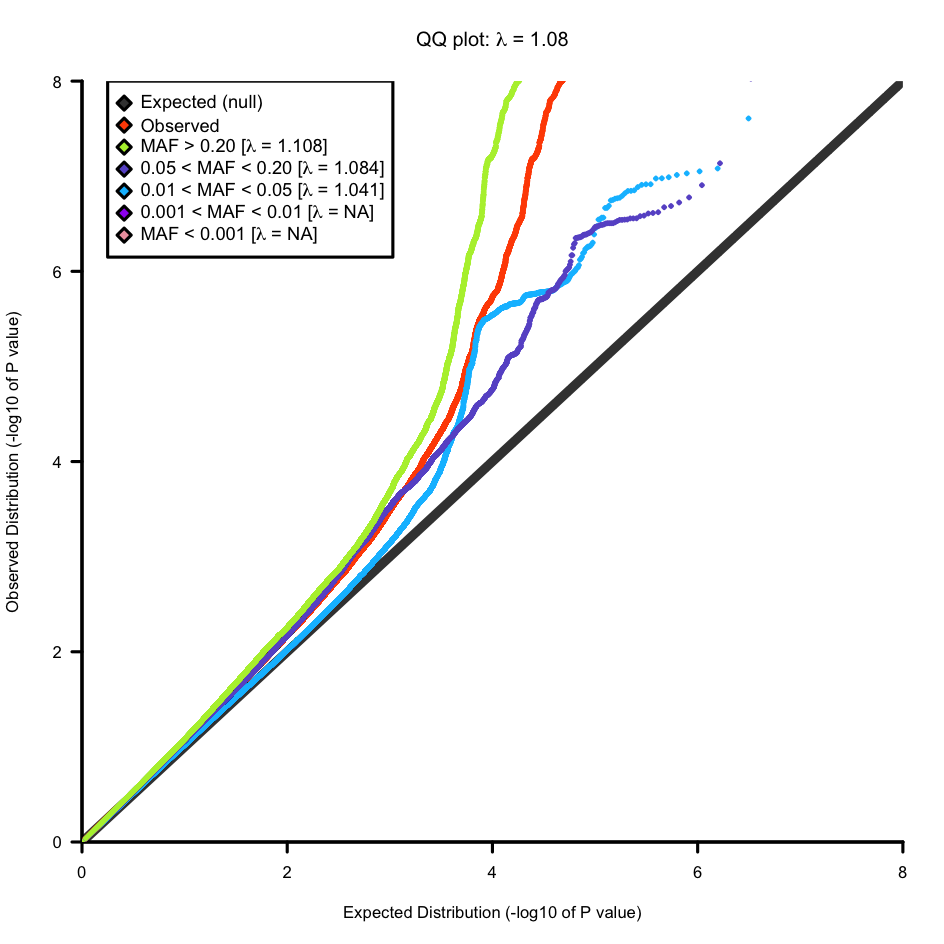

Supplementary Figure 4. QQ plot for alpha angle GWAS meta-analysis

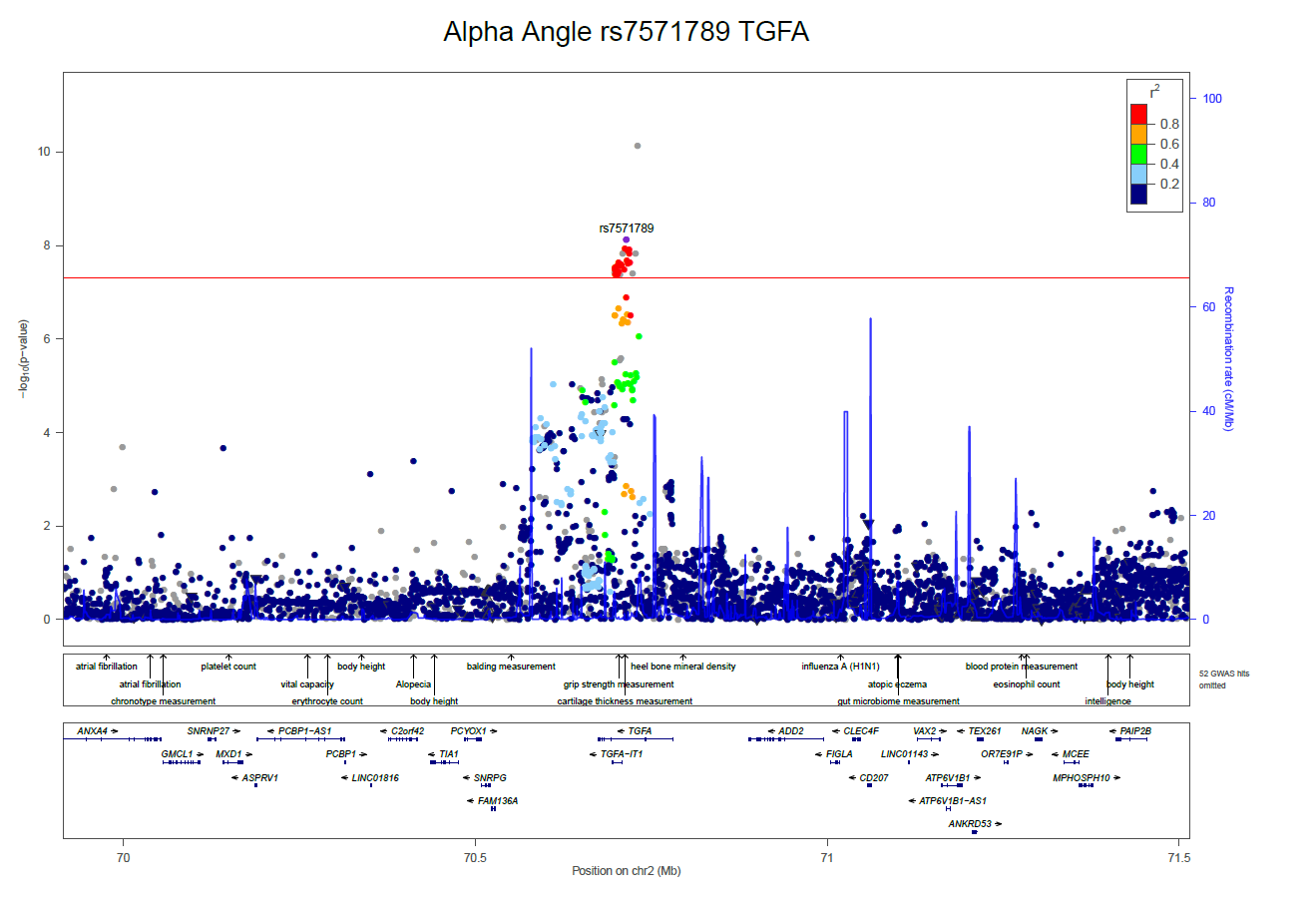

Supplementary Figure 5a. Locus zoom plot rs7571789 (*TGFA*)

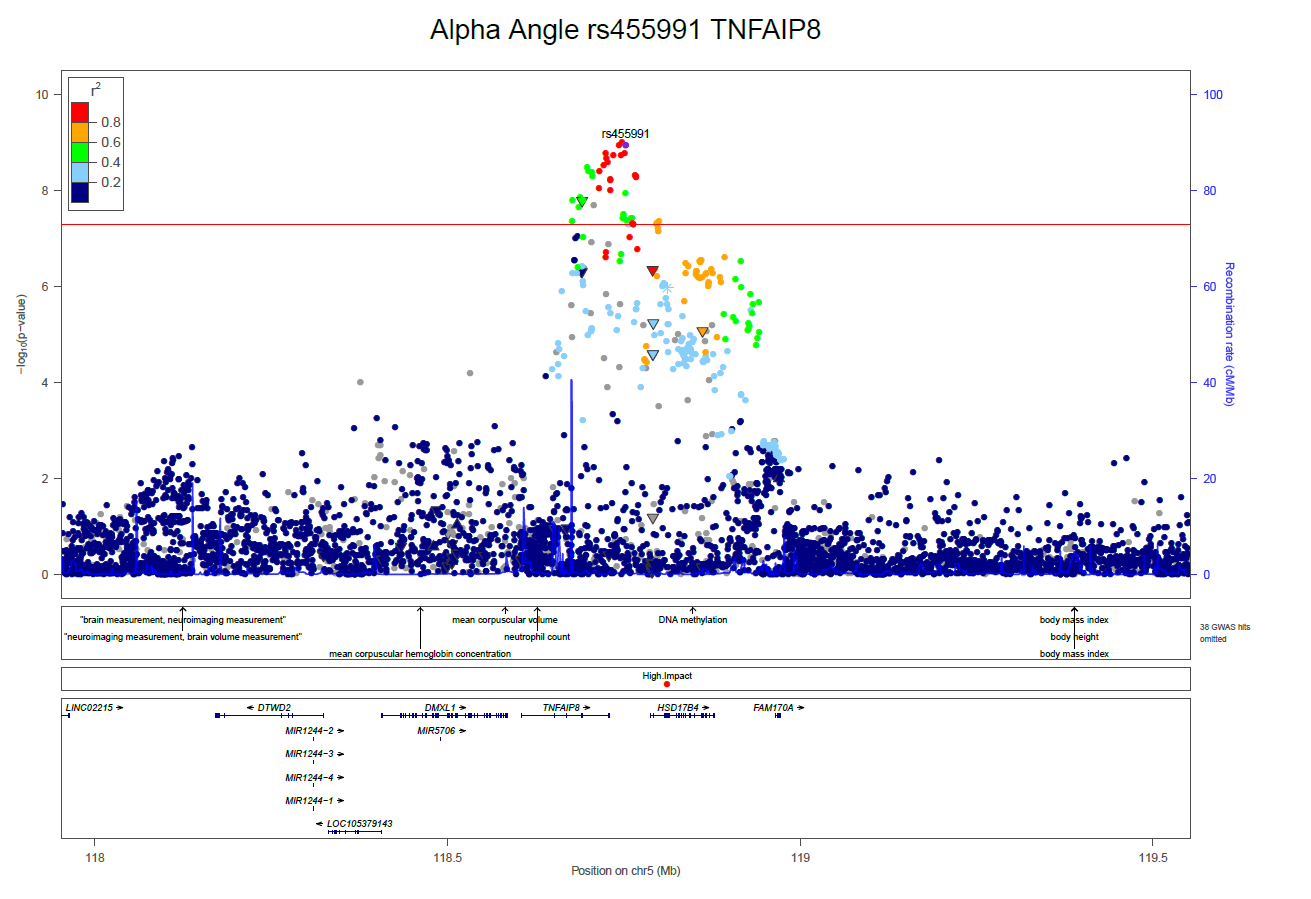

Supplementary Figure 5b. Locus zoom plot rs455991 (*TNFAIP8*)

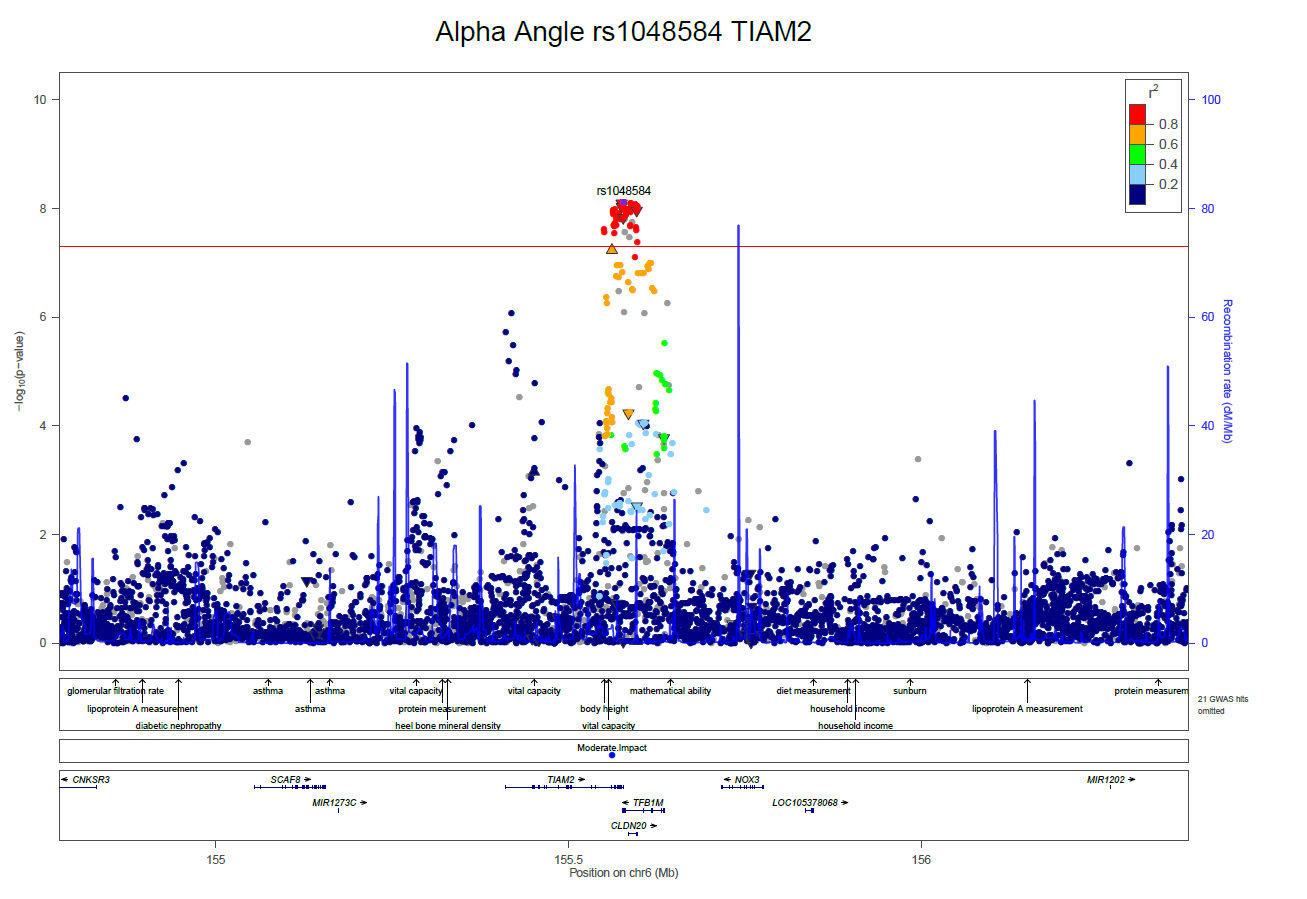

Supplementary Figure 5c. Locus zoom plot rs1048584 (*TIAM2-TFB1M*)

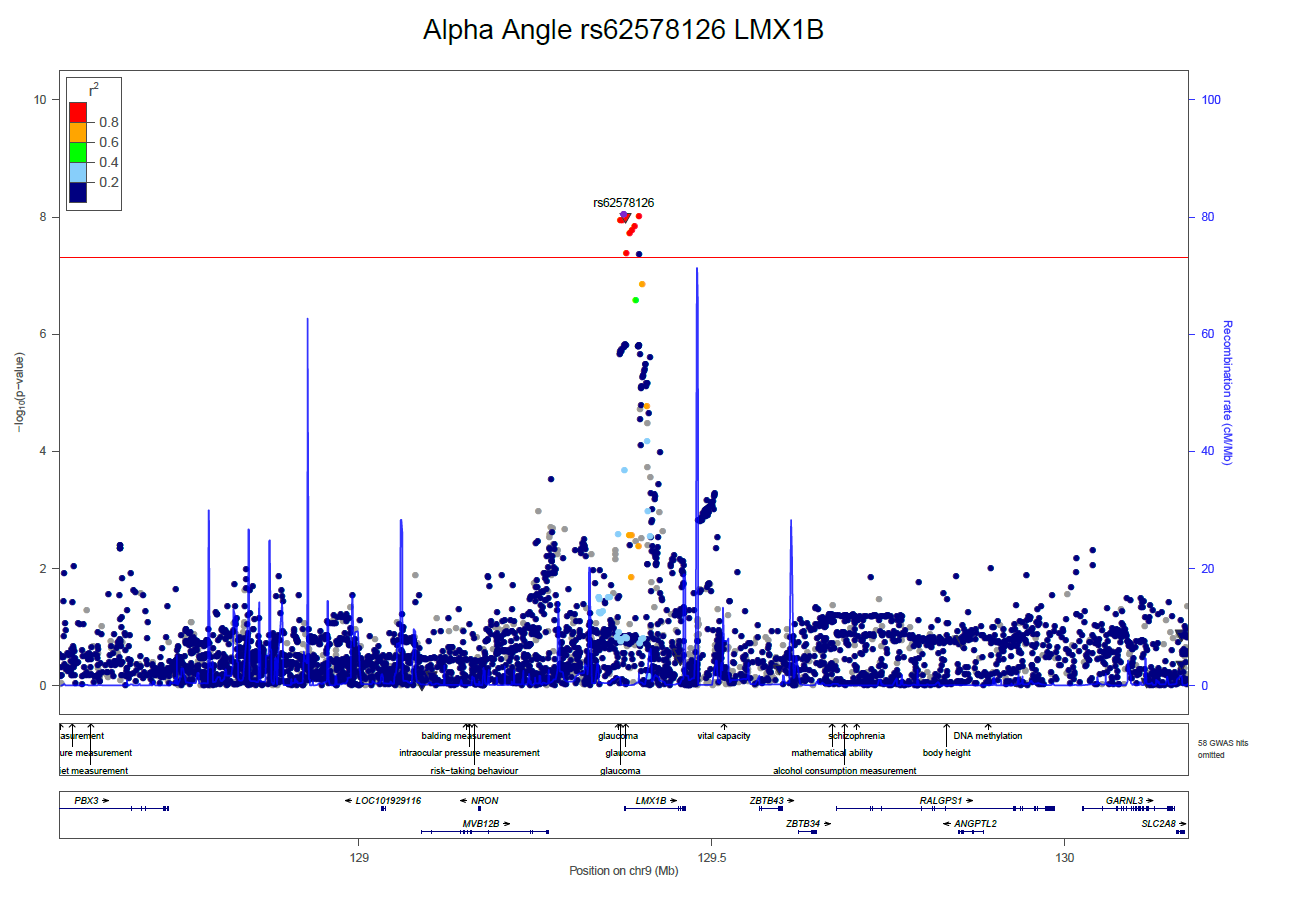

Supplementary Figure 5d. Locus zoom plot rs62578126 (*LMX1B*)

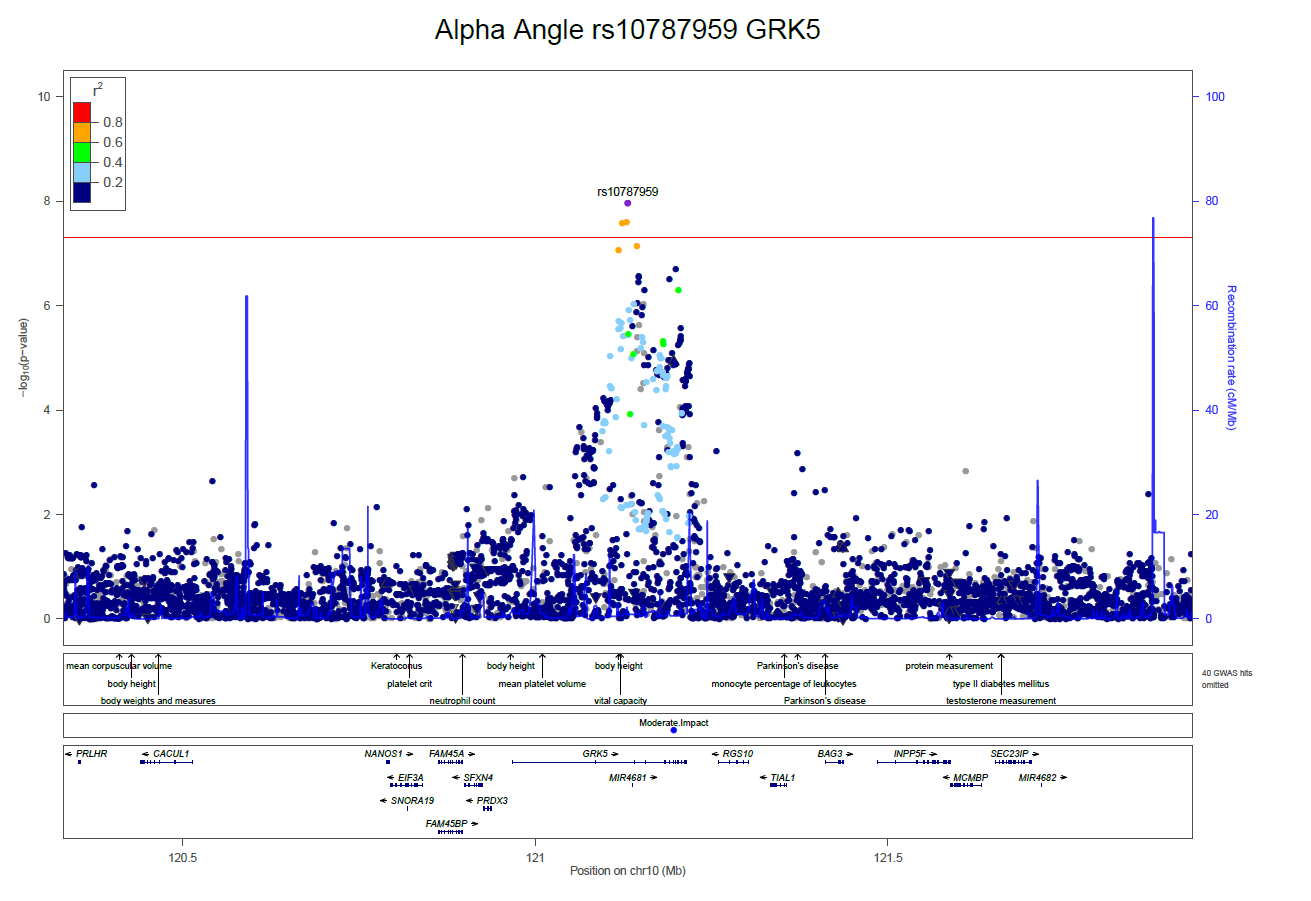

Supplementary Figure 5e. Locus zoom plot rs10787959 (*GRK5*)

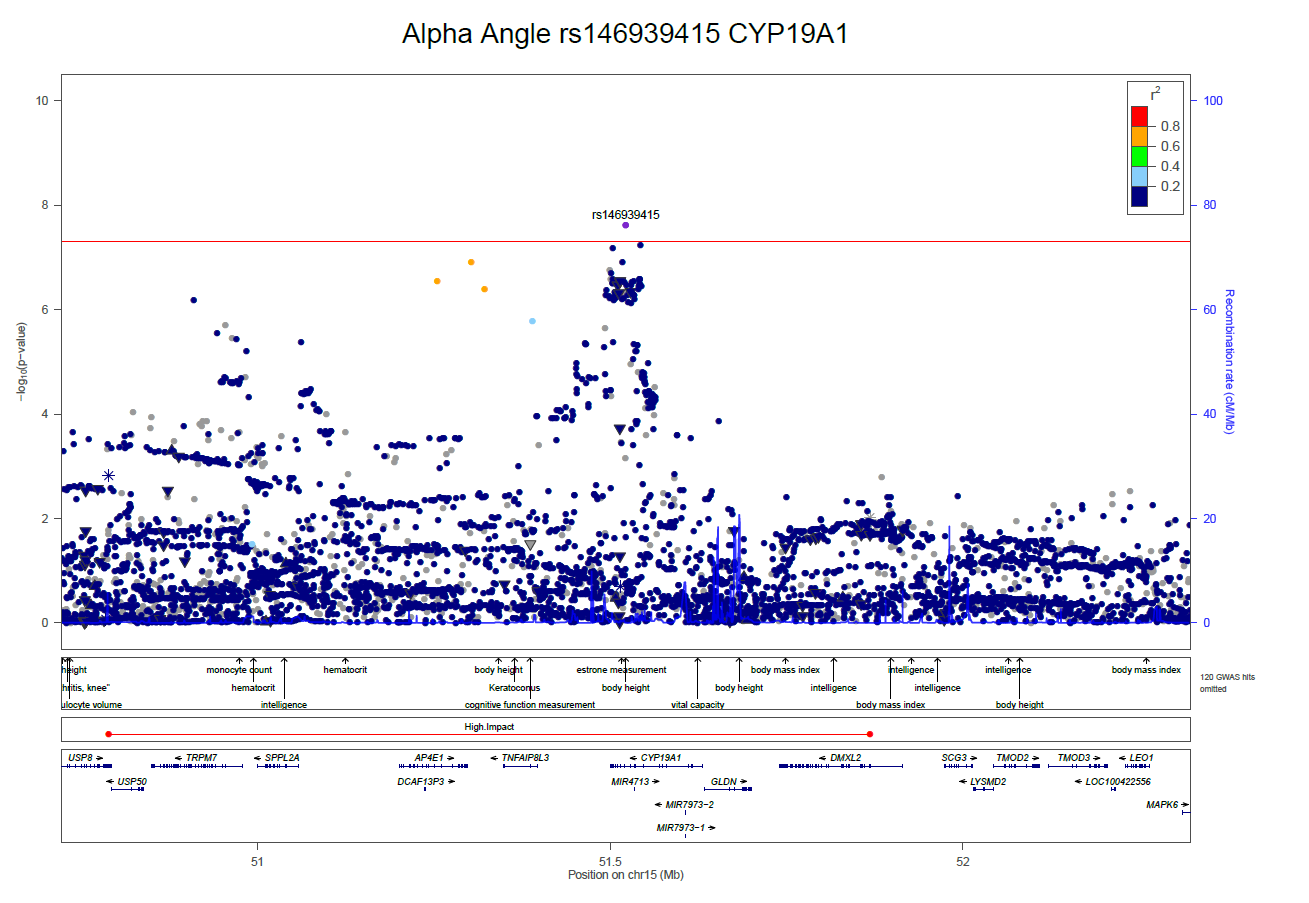

Supplementary Figure 5f. Locus zoom plot rs146939415 (*CYP19A1*)

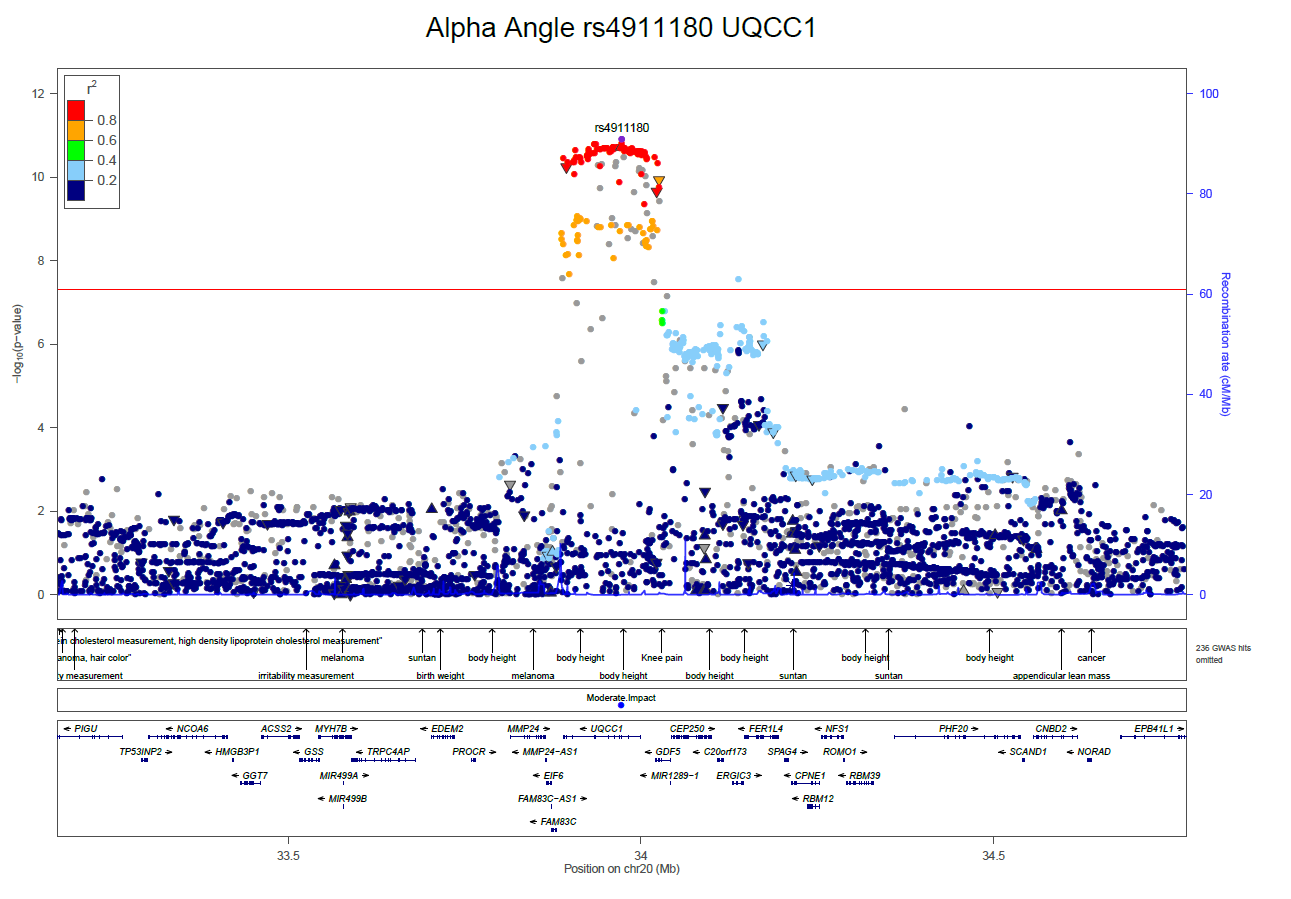

Supplementary Figure 5g. Locus zoom plot rs4911180 (*UQCC1*)

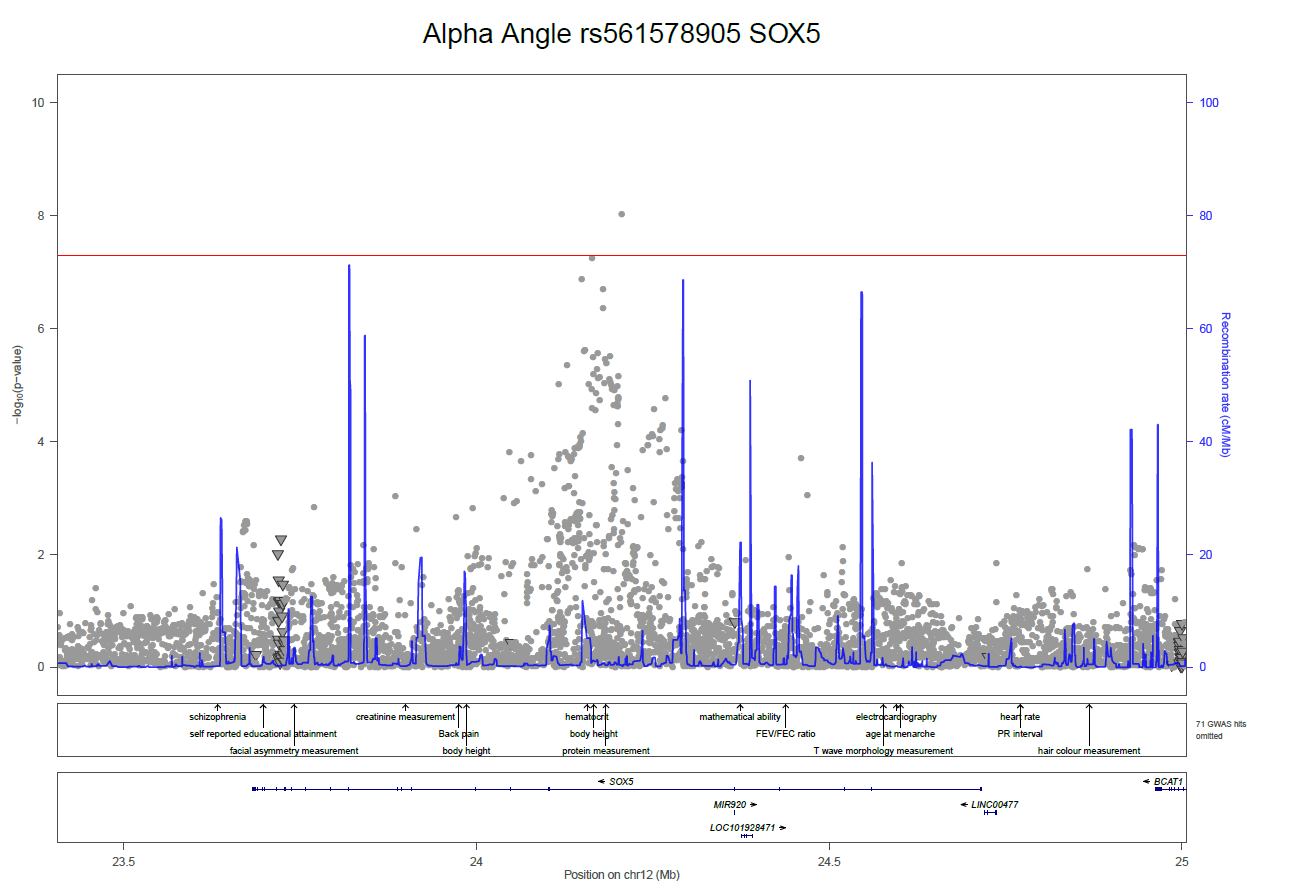

Supplementary Figure 5h. Locus zoom plot rs561578905 (*SOX5*)

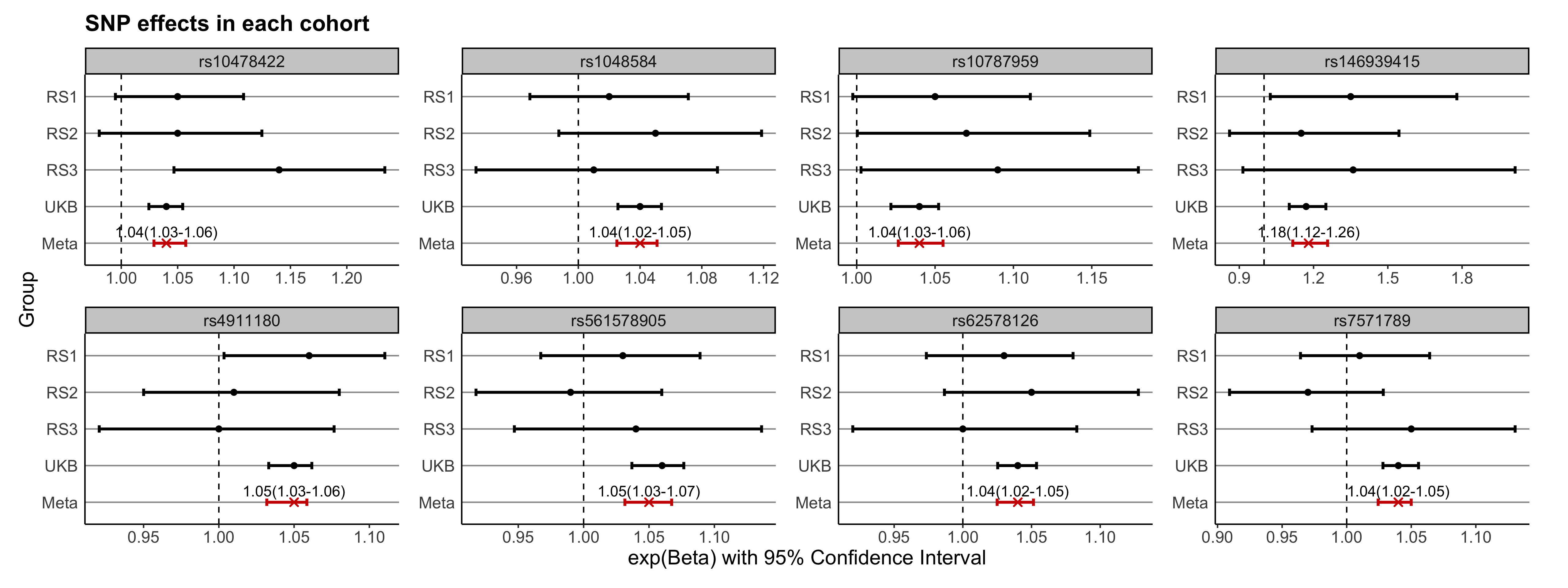

Supplementary Figure 6. A Forest Plot for each independent SNP.

The SNP effects from each cohort are displayed. The exponentiated beta is displayed to aid visualisation. The heterogeneity statistic (I^2^) was zero for all SNPs apart from rs7571789 (I^2^ = 53, P-value = 0.09), rs10478422 (I^2^ = 33, P = 0.21) and rs561578905 (I^2^ = 25, P = 0.26). RS - Rotterdam Study, UKB - UK Biobank, Meta - Meta-analysis.

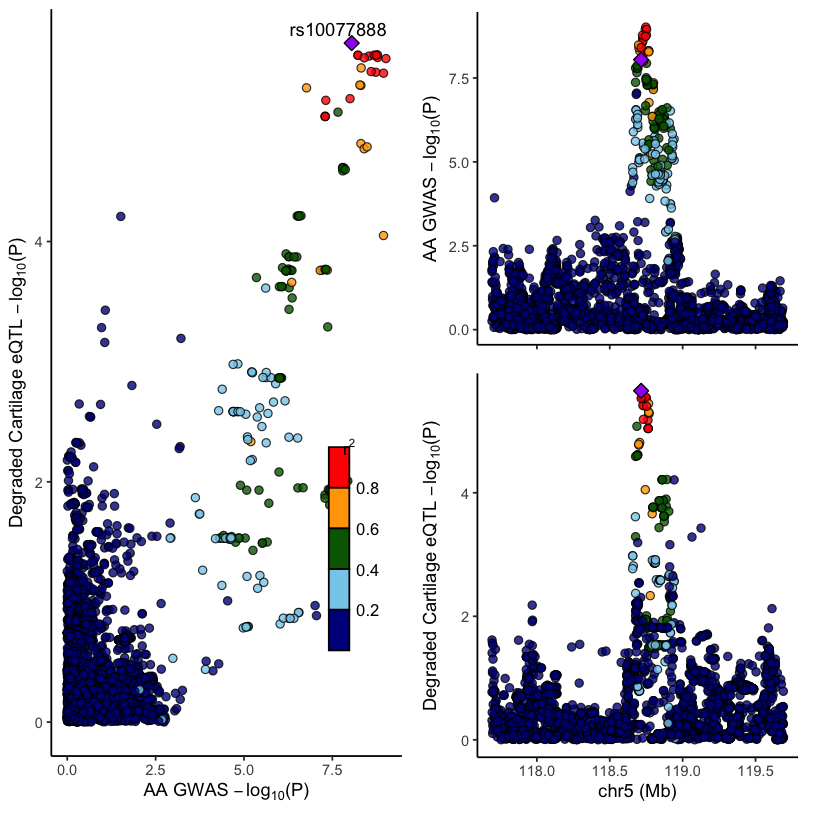

Supplementary Figure 7. A colocalisation plot for *TNFAIP8* expression in highly degraded human cartilage.

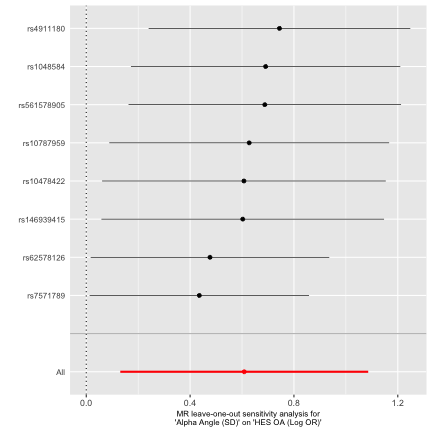

Supplementary Figure 8a. Leave one out analysis comparing alpha angles effect on hip osteoarthritis.

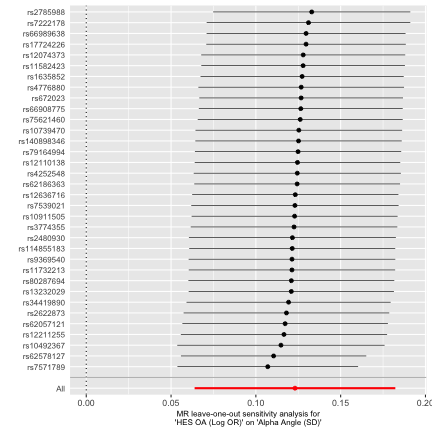

Supplementary Figure 8b. Leave one out analysis comparing hip osteoarthritis effect on alpha angle.

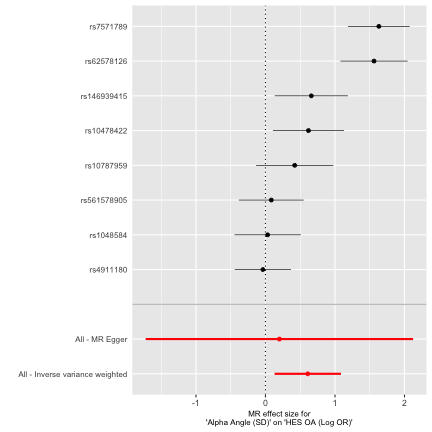

Supplementary Figure 9a. Single SNP analysis of alpha angles effect on hip osteoarthritis.

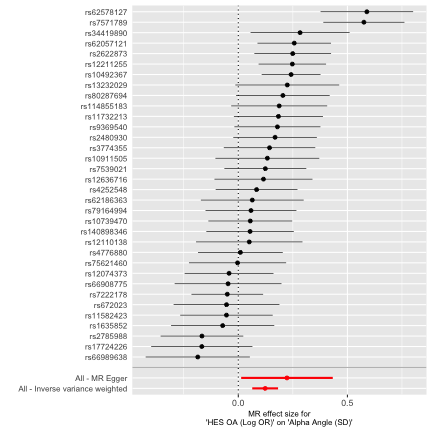

Supplementary Figure 9b. Single SNP analysis of hip osteoarthritis effect on alpha angle.

Refences:

1.	Bycroft C, Freeman C, Petkova D, Band G, Elliott LT, Sharp K, et al. The UK Biobank resource with deep phenotyping and genomic data. Nature. 2018;562(7726):203-9.

2.	Harvey NC, Matthews P, Collins R, Cooper C, Group UKBMA. Osteoporosis epidemiology in UK Biobank: a unique opportunity for international researchers. Osteoporos Int. 2013;24(12):2903-5.

3.	Littlejohns TJ, Holliday J, Gibson LM, Garratt S, Oesingmann N, Alfaro-Almagro F, et al. The UK Biobank imaging enhancement of 100,000 participants: rationale, data collection, management and future directions. Nature Communications. 2020;11(1):2624.

4.	Lindner C, Thiagarajah S, Wilkinson JM, arc OC, Wallis GA, Cootes TF. Development of a fully automatic shape model matching (FASMM) system to derive statistical shape models from radiographs: application to the accurate capture and global representation of proximal femur shape. Osteoarthritis Cartilage. 2013;21(10):1537-44.

5.	Lindner C, Thiagarajah S, Wilkinson JM, arc OC, Wallis GA, Cootes TF. Fully automatic segmentation of the proximal femur using random forest regression voting. IEEE Trans Med Imaging. 2013;32(8):1462-72.

6.	Faber BG, Ebsim R, Saunders FR, Frysz M, Lindner C, Gregory JS, et al. A novel semi-automated classifier of hip osteoarthritis on DXA images shows expected relationships with clinical outcomes in UK Biobank. Rheumatology (Oxford). 2021.

7.	Faber BG, Ebsim R, Saunders FR, Frysz M, Lindner C, Gregory JS, et al. Osteophyte size and location on hip DXA scans are associated with hip pain: findings from a cross sectional study in UK Biobank. Bone. 2021:116146.

8.	Faber B. benfaber20/Automatic-alpha-angle: Alpha Angle from DXA v1.2 (Version v1.2). Zenodo. 2021, January 25;<http://doi.org/10.5281/zenodo.4462770>.

9.	Faber BG, Ebsim R, Saunders FR, Frysz M, Davey Smith G, Cootes T, et al. Deriving alpha angle from anterior-posterior dual-energy x-ray absorptiometry scans: an automated and validated approach. Wellcome Open Research. 2021(<https://wellcomeopenresearch.org/articles/6-60/v1>).

10.	Faber BG, Ebsim R, Saunders FR, Frysz M, Gregory JS, Aspden RM, et al. Cam morphology but neither acetabular dysplasia nor pincer morphology is associated with osteophytosis throughout the hip: findings from a cross-sectional study in UK Biobank. Osteoarthritis Cartilage. 2021;29(11):1521-9.

11.	Zengini E, Hatzikotoulas K, Tachmazidou I, Steinberg J, Hartwig FP, Southam L, et al. Genome-wide analyses using UK Biobank data provide insights into the genetic architecture of osteoarthritis. Nat Genet. 2018;50(4):549-58.

12.	Ikram MA, Brusselle G, Ghanbari M, Goedegebure A, Ikram MK, Kavousi M, et al. Objectives, design and main findings until 2020 from the Rotterdam Study. Eur J Epidemiol. 2020;35(5):483-517.

13.	Agricola R, Heijboer MP, Bierma-Zeinstra SMA, Verhaar JAN, Weinans H, Waarsing JH. Cam impingement causes osteoarthritis of the hip: a nationwide prospective cohort study (CHECK). Annals of the Rheumatic Diseases. 2013;72(6):918-23.

14.	Bycroft C, Freeman C, Petkova D, Band G, Elliott LT, Sharp K, et al. The UK Biobank resource with deep phenotyping and genomic data. Nature. 2018;562(7726):203-9.

15.	Das S, Forer L, Schonherr S, Sidore C, Locke AE, Kwong A, et al. Next-generation genotype imputation service and methods. Nat Genet. 2016;48(10):1284-7.

16.	Abdulrahim H, Jiao Q, Swain S, Sehat K, Sarmanova A, Muir K, et al. Constitutional morphological features and risk of hip osteoarthritis: a case-control study using standard radiographs. Ann Rheum Dis. 2020.

17.	Winkler TW, Day FR, Croteau-Chonka DC, Wood AR, Locke AE, Magi R, et al. Quality control and conduct of genome-wide association meta-analyses. Nat Protoc. 2014;9(5):1192-212.

18.	Boer CG, Hatzikotoulas K, Southam L, Stefansdottir L, Zhang Y, Coutinho de Almeida R, et al. Deciphering osteoarthritis genetics across 826,690 individuals from 9 populations. Cell. 2021;184(18):4784-818 e17.

19.	Trajanoska K, Morris JA, Oei L, Zheng HF, Evans DM, Kiel DP, et al. Assessment of the genetic and clinical determinants of fracture risk: genome wide association and mendelian randomisation study. BMJ. 2018;362:k3225.

20.	Wood AR, Esko T, Yang J, Vedantam S, Pers TH, Gustafsson S, et al. Defining the role of common variation in the genomic and biological architecture of adult human height. Nat Genet. 2014;46(11):1173-86.

21.	Locke AE, Kahali B, Berndt SI, Justice AE, Pers TH, Day FR, et al. Genetic studies of body mass index yield new insights for obesity biology. Nature. 2015;518(7538):197-206.

22.	Morris JA, Kemp JP, Youlten SE, Laurent L, Logan JG, Chai R, et al. An Atlas of Human and Murine Genetic Influences on Osteoporosis. 2018.

23.	Bulik-Sullivan B, Finucane HK, Anttila V, Gusev A, Day FR, Loh PR, et al. An atlas of genetic correlations across human diseases and traits. Nat Genet. 2015;47(11):1236-41.

24.	Brittberg M, Peterson L. Introduction of an articular cartilage classification. ICRS Newsletter. 1998;1(1):5-8.

25.	Peterson L, Minas T, Brittberg M, Nilsson A, Sjogren-Jansson E, Lindahl A. Two- to 9-year outcome after autologous chondrocyte transplantation of the knee. Clin Orthop Relat Res. 2000(374):212-34.

26.	Binch A, Snuggs J, Le Maitre CL. Immunohistochemical analysis of protein expression in formalin fixed paraffin embedded human intervertebral disc tissues. JOR Spine. 2020;3(3):e1098.
